# Associations of Metabolic Vulnerability Index with Cardiometabolic Diseases, Multimorbidity, and All-cause Mortality

**DOI:** 10.64898/2026.02.09.26345939

**Authors:** Shiyu Zhu, Gaoyu Yu, Yunrui Lu, Hui Ni, Jian Shen, Chaoyue Zhao, Yue Zhang, Xiaolin Xu, Meixiang Xiang, Yao Xie

## Abstract

**Background:** Metabolic vulnerability index (MVX), a novel biomarker of systemic inflammation and metabolic malnutrition, is associated with mortality in patients with cardiovascular diseases. Nevertheless, little is known about its association with cardiometabolic diseases (CMDs) and multimorbidity (CMM). We aimed to examine the associations of MVX with the risk of individual CMDs, their progression to CMM, and all-cause mortality in the general population.

**Methods:** In a prospective cohort of 218,635 UK Biobank participants free of CMDs, MVX was calculated based on plasma metabolomics data. CMM was defined as coexistence of two or more CMDs, including coronary heart disease (CHD), stroke, and type 2 diabetes mellitus (T2DM). Cox proportional hazard and multi-state models were employed to evaluate the associations of MVX with the risks of individual CMDs, CMM, and all-cause mortality.

**Results:** During a median follow-up of 14.4 years, 27,805 (12.7%) participants developed at least one CMD, 3,006 (1.4%) progressed to CMM, and 14,211 (6.5%) died. Each standard deviation increase of MVX score was associated with 9% (95% confidence interval: 8%-10%), 11% (7%-15%), and 12% (10%-14%) higher risks of developing CMDs, CMM, and mortality, respectively. The MVX-CMM associations were more prominent in females and in the sequential onset pattern of T2DM followed by CHD or stroke. Multi-state model analysis further uncovered consistent associations between higher scores of MVX and higher risks of transitions from CMDs free to CMD, subsequently to CMM, and to death.

**Conclusions:** Higher MVX scores were significantly associated with higher risks of incident CMDs, their progressions to CMM, and all-cause mortality. These results underscored the potential of MVX in the primary prevention and management of CMDs and CMM.

## 1 Introduction

Cardiometabolic multimorbidity (CMM) refers to the coexistence of two or more cardiometabolic diseases (CMDs) including coronary heart disease (CHD), stroke, and type 2 diabetes mellitus (T2DM) [1]. In the context of population aging, the prevalence of CMDs and CMM is progressively increasing which imposes a great health burden [2]. Compared to patients with individual CMDs, those with CMM were associated with worsened life quality and shorter life expectancy [3–5]. Inflammation and malnutrition play an important role in the pathophysiology of CMDs and CMM [6, 7]. Systemic inflammation is a key driver of atherogenesis and maladaptive cardiovascular remodeling [8], and links the association between adiposity and insulin resistance [9]. Metabolic malnutrition fosters inflammation, sarcopenia, and frailty, resulting in elevated incidence and deteriorated prognosis of various chronic diseases [10]. However, there is still an unmet need for precise phenotyping of nutritional and inflammatory status to refine the prevention and management of CMDs and CMM.

Metabolomics serves as a powerful tool for risk evaluation of CMDs [11]. The metabolic vulnerability index (MVX) is a novel biomarker reflecting systemic inflammation and metabolic malnutrition of an individual, which is constructed based on metabolomics data generated by plasma nuclear magnetic resonance (NMR) spectroscopy [12]. Initially developed for the prediction of mortality in people at a high risk of CHD [12], MVX also improved the classification of mortality risk beyond existing well-established clinical markers in patients with heart failure (HF) [13]. A recent study further illustrated significant associations between MVX and mortality in a prospective cohort [14]. Considering the role of inflammation and malnutrition in CMDs, we hypothesized potential associations between MVX, CMDs, and CMM, which have not been investigated in the general population without known CMDs. Evidence for the association of MVX with the incidence and progression of CMDs and CMM would broaden the application spectrum of MVX from secondary prevention to primary prevention.

Herein, we aim to assess the associations of MVX with the incidence of individual CMDs, and their progressions to CMM and mortality in a prospective cohort of UK Biobank (UKB). Multi-state model (MSM) was further employed to explore the potentially heterogeneous associations of MVX score with the disease transitions from healthy status to individual CMD, and subsequent CMM and death.

## 2 Methods

### 2.1 Study population

This study was conducted based on a population from UKB, which is a large-scale population-based cohort recruiting over half a million community-dwelling middle-aged and older adults between 2006 and 2010 [15]. Sociodemographic characteristics, lifestyles factors, and blood samples were collected at baseline using touchscreen questionnaires, and health-related outcomes were followed through the linkage to national health registers. UKB received ethical approval from the North West Multicenter Research Ethics Committee (reference number 11/NW/0382) and all participants have given written informed consents. This study has been approved by the UKB Access Management Team according to established access procedures (Application ID: 83421).

We included all participants with available NMR metabolomics data as the initial study population (*n* = 274,351). Participants with CHD, stroke, or diabetes at baseline (*n* = 15,628, 5,192, and 17,242, respectively) were excluded. Furthermore, we excluded participants with cancer (*n* = 24,893) in consistent with previous studies of CMM [16, 17]. We also excluded participants who had missing values in any component of MVX (*n* = 259), or those who withdrew consent or were lost to follow-up (*n* = 1,282). Finally, a total of 218,635 participants were included in the analysis. The flowchart of the study is depicted in **Figure 1A**, and detailed exclusion criteria are shown in **Additional File 1: Table S1**.

**Figure 1.**
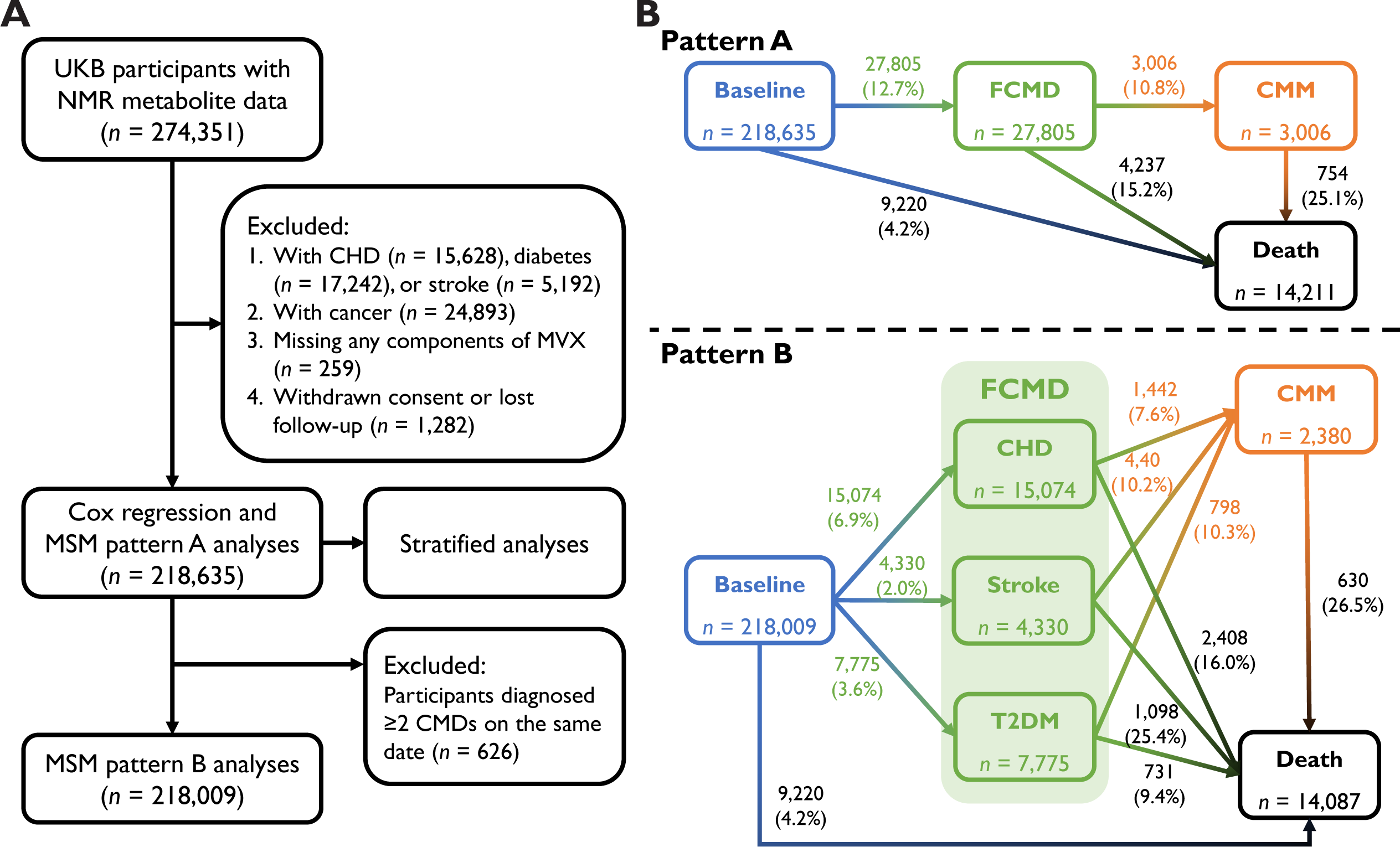
Flow chart of the study and transition patterns of the multi-state model analysis. (A) Flow chart of the study. Some excluded participants met two or more exclusion criteria simultaneously. (B) Number (percentages) of participants in each transition stage in the MSM analysis. NMR, nuclear magnetic resonance; MVX, metabolic vulnerability index; MSM, multi-state model; CMD, cardiometabolic disease; FCMD, first cardiometabolic disease; CMM, cardiometabolic multimorbidity; CHD, coronary heart disease; T2DM, type 2 diabetes mellitus.

### 2.2 Assessment of MVX

Metabolomics data were measured using a high-throughput NMR-based platform developed by Nightingale Health based on ethylenediaminetetraacetic acid (EDTA) plasma samples collected at the baseline assessment [18, 19]. Sex-specific MVX scores were calculated based on NMR-metabolite data as previously described [12] (**Table 1**). Briefly, two biomarkers of systemic inflammation, glycoprotein acetyls (GlycA) and small high-density lipoprotein particles (S-HDLP) [20], were combined into the inflammation vulnerability index (IVX). The metabolic malnutrition index (MMX), which reflects protein-energy wasting, was calculated through three branched chain amino acids (leucine, isoleucine, and valine) and citrate. Finally, IVX and MMX were incorporated into MVX with a score ranging from 1 to 100, which was parallel to the level of systemic inflammation and protein-energy wasting [12]. MVX score was modelled as both continuous variable (per SD increase) and sex-specific quartiles.

**Table 1.**
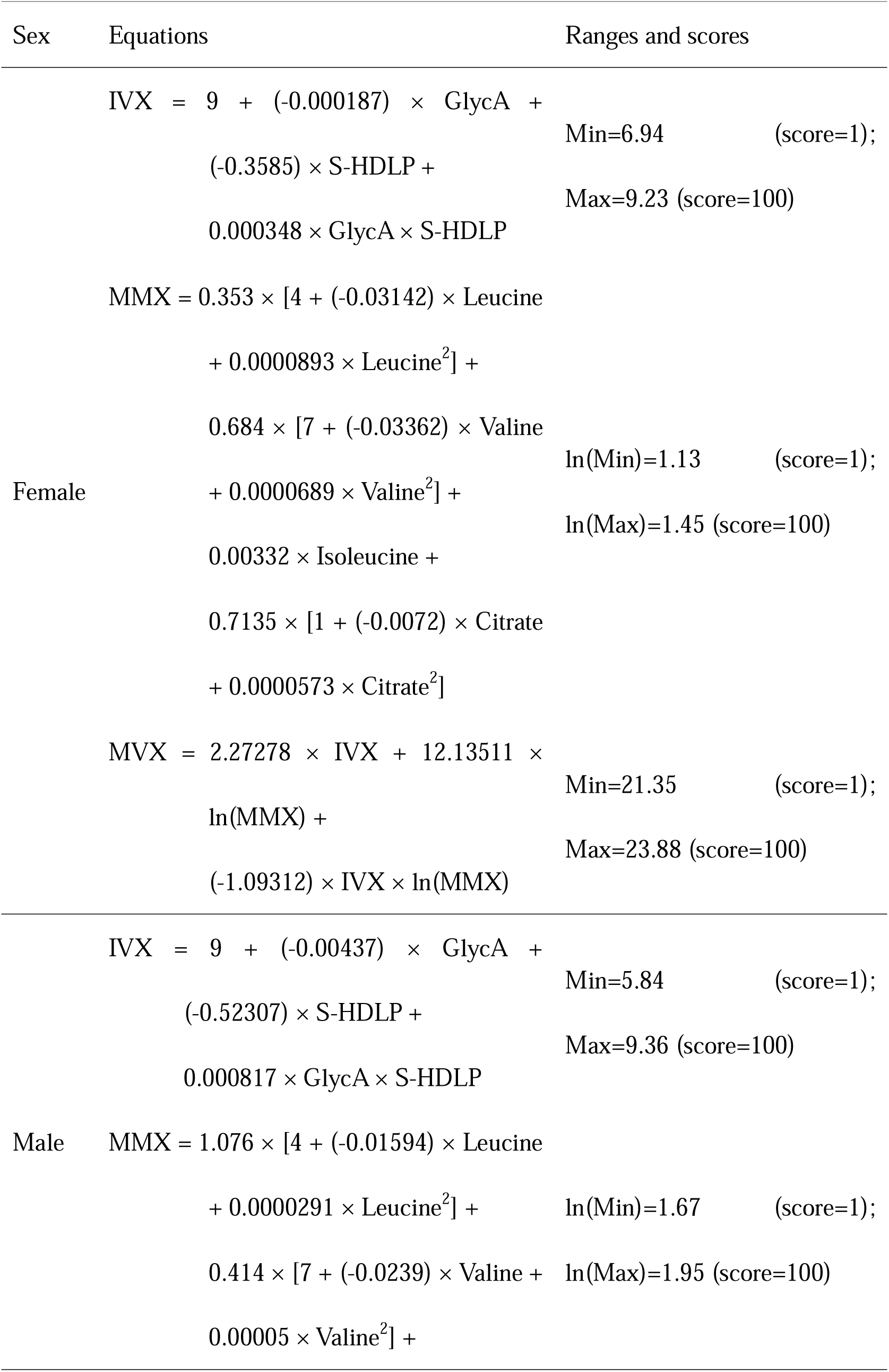
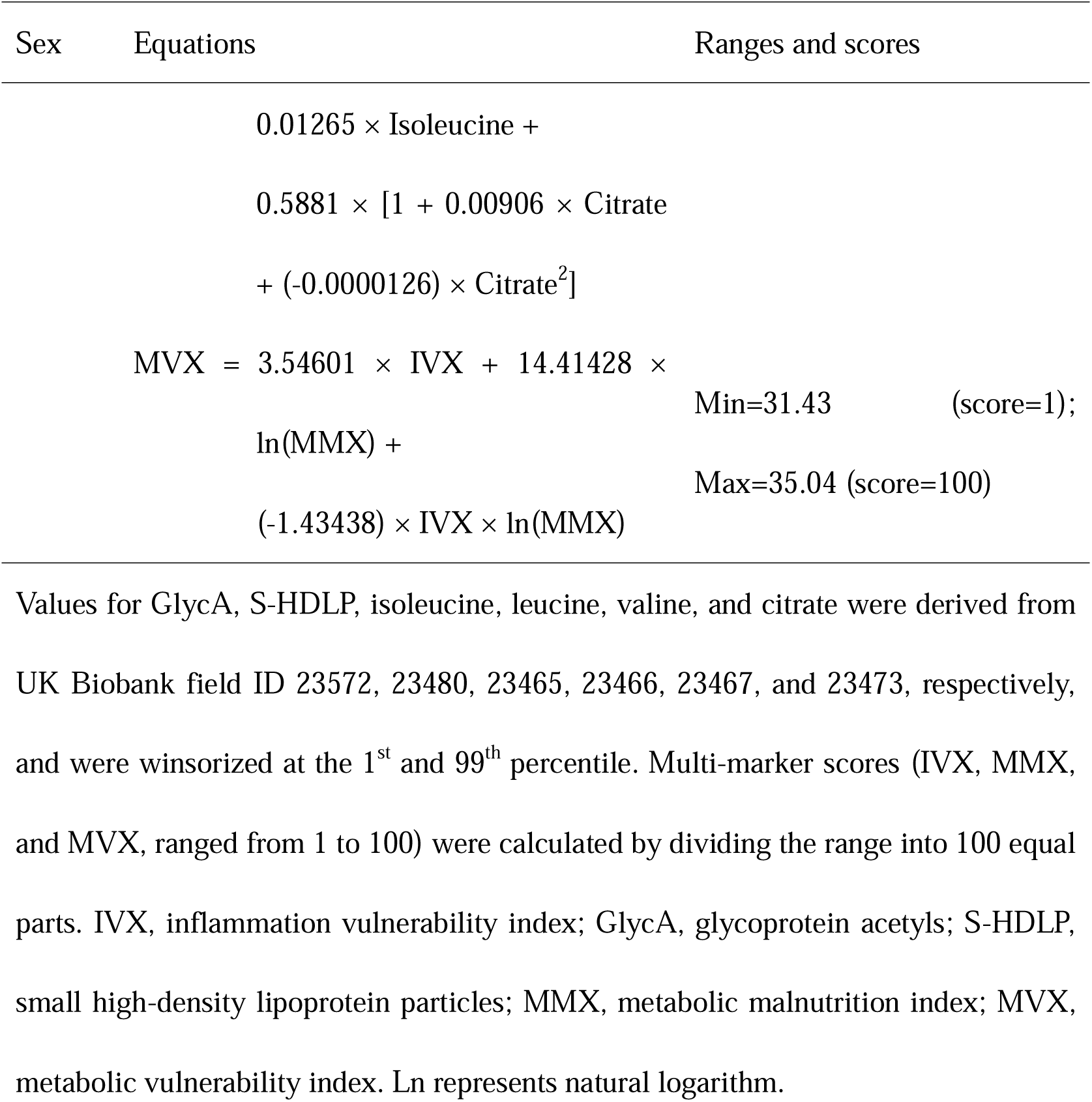
Calculation of MVX score in the UKB cohort.

### 2.3 Ascertainment of outcomes

The primary outcome of the study was the incidence of individual CMDs, their progressions to CMM, and all-cause mortality. In line with previous studies, we limited CMDs to CHD, stroke, and T2DM [16, 21]. Incident CMDs were ascertained based on linkage with primary care, hospital inpatient records, and death register. Following previous literature, CHD, stroke, and T2DM were coded as the ICD-10 codes I20-I25, I60-I64 and I69, and E11 and E14, respectively [16, 17]. As the majority of UKB participants were middle- or old-aged participants, it was reasonable to assume that unspecified diabetes (E14) were mostly T2DM. CMM was defined as the coexistence of two or more CMDs in an individual. Information on all-cause mortality was identified through the linkage to national death registries. Participants were censored at death or the end of follow-up (July 1^st^, 2023), whichever came first. Detailed definitions of study outcomes are available in **Additional File 1: Table S2**.

### 2.4 Covariates

To reduce the potential confounding, a series of covariates including socio-demographic characteristics, lifestyle factors, medical conditions, and familial history were selected according to empirical evidence [16, 21] and *a priori* defined directed acyclic graph (DAG, **Additional File 1: Figure S1**). Detailed source and definitions of covariates are shown in **Additional File 1: Table S3**. Missing covariates were imputed with sex-specific median for continuous variables and sex-specific mode for categorical variables in the main analysis. We also utilized multiple imputations by chained equations in sensitivity analysis. The numbers and percentages of missing values for each variable are illustrated in **Additional File 1: Figure S2**.

### 2.5 Statistical analysis

Baseline characteristics of study participants were summarized as mean (standard deviation, SD) or median (interquartile range, IQR) for continuous variables and frequency (percentage) for categorical variables. Pearson or Spearman correlation analyses were utilized to explore potential associations between MVX and covariates. Kaplan-Meier curves and log-rank tests were utilized to compare the incidence of individual CMDs, CMM, and mortality across different groups of MVX. In the main analysis, we used Cox proportional hazards models to assess the associations of MVX and its components with the risks of individual CMDs, CMM, and all-cause mortality. The fully adjusted model included age at baseline, sex, ethnicity, educational level, Townsend deprivation index, smoking status, drinking status, ideal diet as previous described [22], sleep duration, physical activity, body mass index (BMI), parental history of CMM, history of hypertension and dyslipidemia, and hemoglobin A1c (HbA1c) levels (**Additional File 1: Table S3**). Additionally, restricted cubic spline analysis was performed with four knots at 5^th^, 35^th^, 65^th^ and 95^th^ percentiles. The discrimination ability of MVX was assessed using Uno’s C-index, integrated discrimination improvement (IDI), and net reclassification improvement (NRI).

We further fitted multinomial logistics regression models to assess the associations between MVX and sequential onset pattern of CMDs [23], i.e., vascular diseases (CHD or stroke) after T2DM and *vice versa*.

To evaluate the association between MVX and the temporal progression of CMM, we conducted unidirectional multi-state models (MSM) analysis [24]. MSM is an advanced derivative of the competing risk model proposed by Fine and Gray, which offers a unique advantage in investigating potential heterogeneity across different stages of disease progression [25]. Since the strict Markov assumption is commonly violated in the settings of chronic diseases (formally evaluated in **Additional File 1: Table S4**), we employed the “clock-reset” (also known as the semi-Markov) approach, rather than the standard “clock-forward” (Markov) approach [26, 27]. Five transitions were considered based on the natural progression trajectory of CMM [16, 17]: (1) from healthy state to the first cardiometabolic disease (FCMD); (2) from healthy state to death without CMDs; (3) from FCMD to CMM; (4) from FCMD to death without CMM; (5) from CMM to death (**Figure 1B**, transition pattern A). For participants entered multiple stages simultaneously (*n* = 1,251), the entrance date of the prior state was calculated as the entrance date of the subsequent state minus 0.5 day [16, 17]. For example, if someone was diagnosed with stroke and died on the same date, the entrance date of FCMD was calculated as the death date minus 0.5 day. We also built MSM separately for CHD, stroke, and T2DM, resulting in 11 transitions (**Figure 1B**, transition pattern B). Participants diagnosed with two or more CMDs simultaneously (*n* = 626) were excluded in the pattern B analysis (**Figure 1A**) due to the uncertainty of the disease onset sequence.

Since MVX is a metabolomic measurement subject to temporal fluctuations, relying solely on baseline values could introduce regression dilution bias. To address this, we analyzed a subsample of 13,707 participants with repeated MVX measurements from 2012 to 2013. The regression dilution ratio (RDR) was calculated by regressing the repeat measurements on the baseline values, adjusted for the interval time, and the corrected log hazard ratios (HRs) and their standard errors were obtained by dividing the observed values by the RDR [28].

We performed stratified analysis according to age at baseline, sex, ethnicity, smoking status, and history of hypertension or dyslipidemia. Potential effect modification of the stratification variables was explored by the Z-test proposed by Altman and Bland [29]. A series of sensitivity analyses were conducted to confirm the robustness of the main results.1) applying several statistical methods to examine the influence of participants who entered multiple stages simultaneously (*n* = 1,251): i) excluding those participants; ii) assigning the entrance date of the prior state as the median transition interval time [30, 31] (1.97 years from FCMD to CMM, 0.31 year from FCMD to death, and 0.25 year from CMM to death); iii) adding another transition from healthy state directly to CMM; 2) redefining T2DM as E11 rather than E11 and E14 to avoid the influence of unspecified diabetes; 3) excluding participants with events occurring during the first two years of follow-up (*n* = 2,923) to avoid reverse causality; 4) additionally adjusted for albumin, CRP, as well as self-reported use of anti-hypertensive medication and cholesterol-lowering medication; 5) excluding participants with missing values in covariates; 6) imputing missing data with multiple imputation by chained equations; 7) including participants with cancer (*n* = 24,893) at baseline; 8) including participants with CMDs at baseline (*n* = 29,893) and assigning them to FCMD or CMM status based on their initial CMD status; 9) additionally adjust for use of statins, and perform stratified analysis by the use of statins; 10) additionally adjust for serum phosphate, an important risk factor of various chronic diseases and mortality [32].

All statistical analyses were performed in R (version 4.6.1). The “mstate” package (version 0.3.3) was utilized for the MSM analysis, and the “survIDINRI” package (version 1.1-2) was used for calculating the IDI and NRI indices. A two-tailed *P*-value <0.05 was considered statistically significant.

## 3 Results

### 3.1 Baseline characteristics of study participants

Among the 218,635 included participants, MVX scores were normally distributed with a mean value of 41.3 (SD: 14.8, **Additional File 1: Figure S3**). The cohort mainly consisted of middle-aged and older white adults with a female proportion of 55.1%. Baseline characteristics stratified by sex-specific quartiles of MVX are summarized in **Additional File 1: Table S5**. Participants with higher MVX scores were more likely to be older, white, smokers, non-drinkers, and tended to have hypertension and dyslipidemia at baseline.

Results of the correlation analysis are illustrated in **Additional File 1: Figure S4**. MVX was positively correlated with several cardiometabolic risk factor such as BMI, CRP, HbA1c, low-density lipoprotein-cholesterol (LDL-C), and triglyceride-glucose (TyG) index, a surrogate marker of insulin resistance [33].

### 3.2 MVX and the risks of individual CMDs, CMM, and all-cause mortality

During a median follow-up of 14.4 years [3.08 million person-years (PYs)], 27,805 (12.7%) participants developed at least one CMD (90.3/10,000 PYs), where 16,615 (7.6%), 5,143 (2.4%), 9,120 (4.2%) participants were diagnosed with CHD, stroke, and T2DM, respectively. Among those who developed at least one CMD, 3,006 (1.4%) further developed CMM (196.4/10,000 PYs). 9,220 (4.2%), 4,237 (15.2%), and 754 (25.1%) participants died without CMDs, with single CMD, and with CMM (29.9/10,000 PYs, 276.9/10,000 PYs, and 618.7/10,000 PYs), respectively (**Figure 1B**). Participants with higher MVX scores had a higher cumulative incidence of individual CMDs, CMM, and all-cause mortality (**Figure 2**).

**Figure 2.**
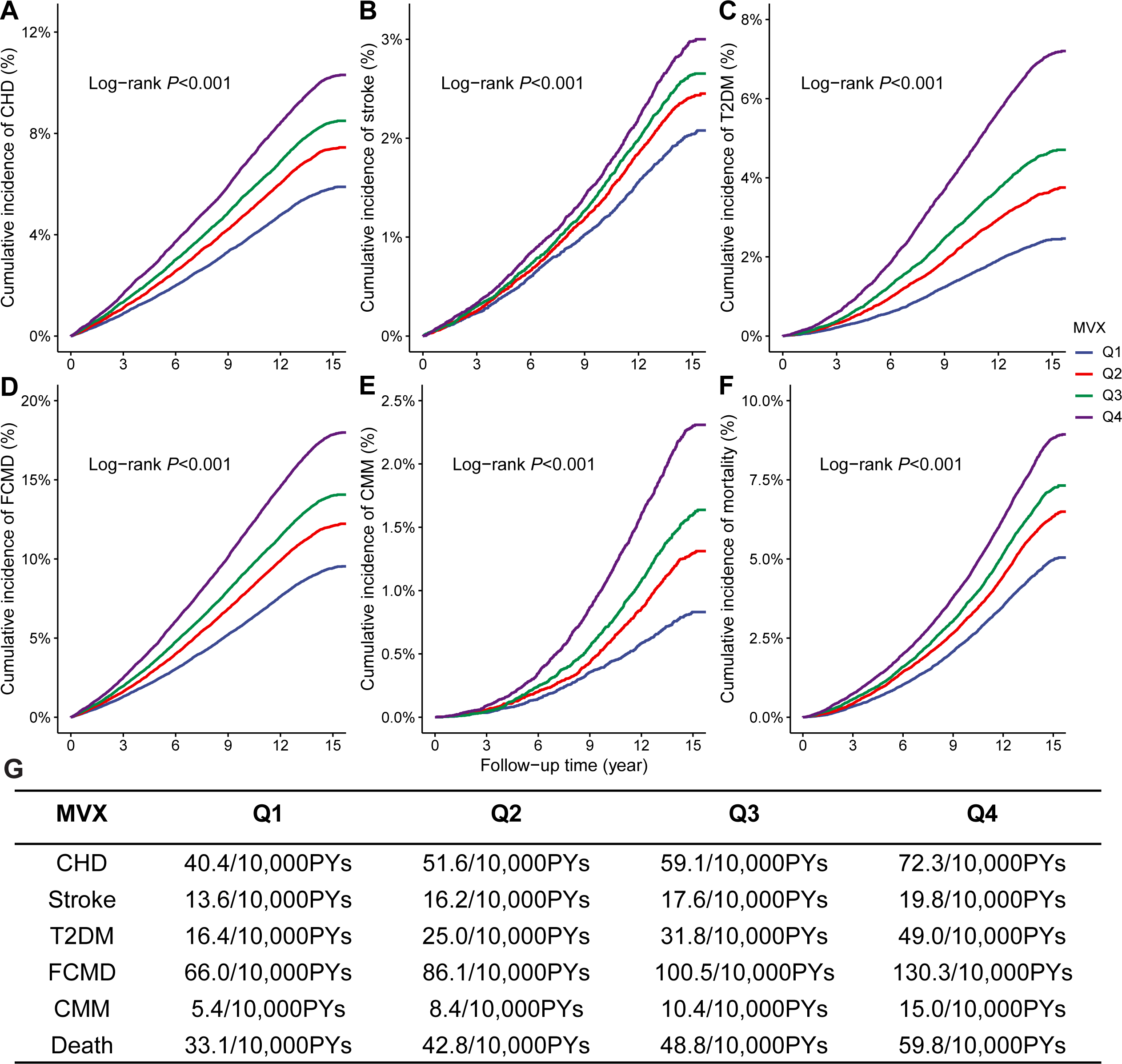
Kaplan-Meier curves for CMDs, CMM, and mortality stratified by sex-specific quartiles of MVX. (A-F) Kaplan-Meier curves of CHD (A), stroke (B), T2DM (C), FCMD (D), CMM (E), all-cause mortality (F) stratified by sex-specific quartiles of MVX. (G) Crude incidence of each outcome per 10,000 person-years (PYs) of follow-up. MVX, metabolic vulnerability index; FCMD, first cardiometabolic disease; CMM, cardiometabolic multimorbidity; CHD, coronary heart disease; T2DM, type 2 diabetes mellitus.

In the multivariable-adjusted Cox regression models (**Figure 3**), each SD increase of MVX was associated with 11% (95%CI: 9%-13%), 4% (1%-7%), 9% (7%-12%), 9% (8%-10%), 11% (7%-15%), and 12% (10%-14%) higher risks of CHD, stroke, T2DM, CMM, and mortality, respectively. Participants with an MVX score in the highest sex-specific quartile group had 1.22-fold (95% CI: 1.17-1.26), 1.35-fold (1.20-1.51), and 1.32-fold (1.26-1.39) higher risks of FCMD, CMM, and all-cause mortality, compared to those in the lowest quartile group (**Additional File 1: Table S6**). The associations between individual MVX components and CMDs, CMM, and mortality are shown in **Additional File 1: Table S7**. For the discrimination ability of MVX, although the absolute increase of C-index seems modest, IDI and NRI demonstrated incremental predictive value of MVX in most cardiometabolic and mortality outcomes (**Additional File 1: Table S8**).

**Figure 3.**
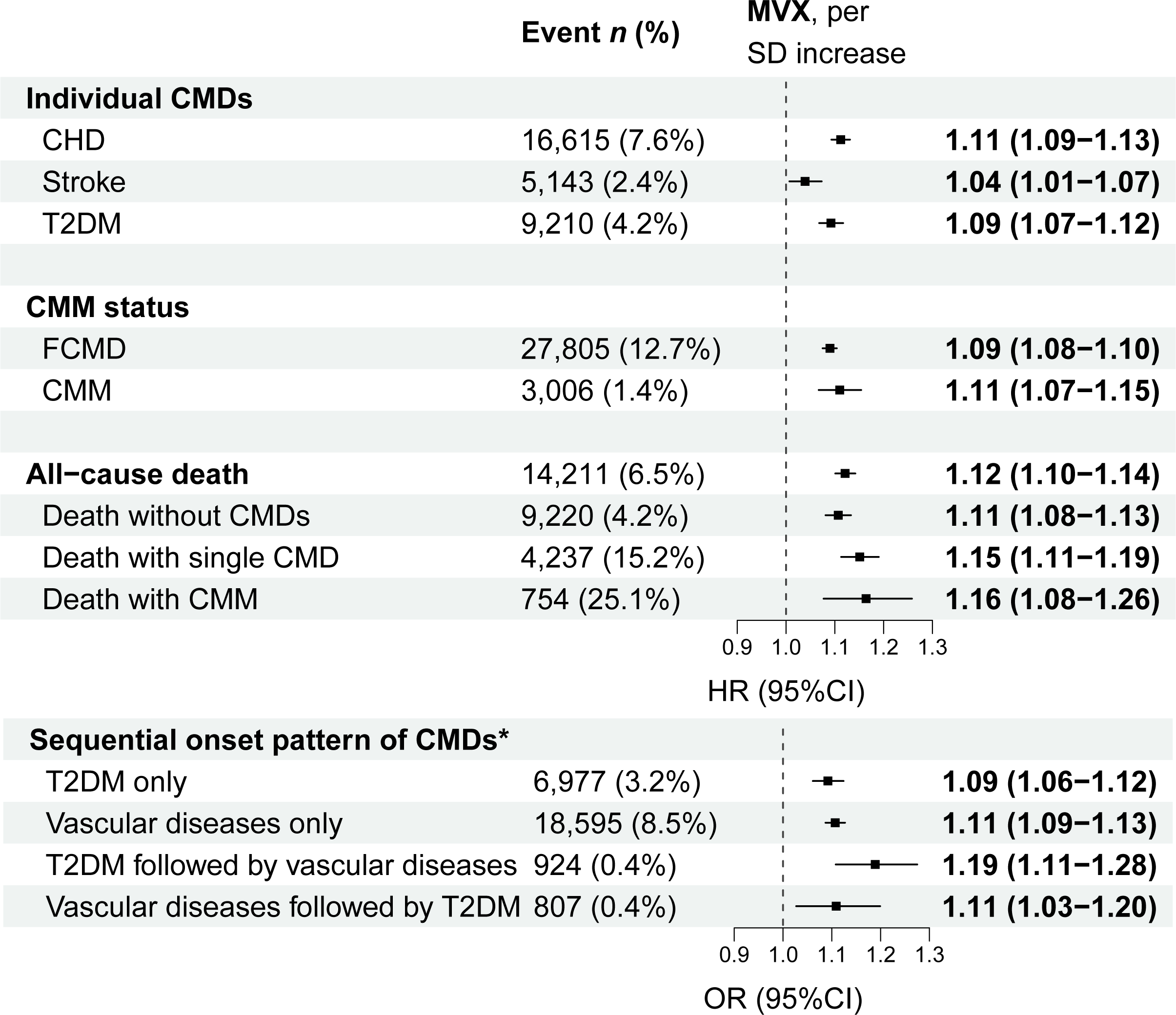
Association between MVX score and incident CMDs, CMM, sequential onset patterns of CMDs, and mortality. Results are HRs (95%CIs) for the MVX-CMDs/CMM/mortality association and ORs (95%CIs) for the MVX-sequential onset pattern associations, per SD increase of MVX, adjusted for age, sex, ethnicity, education, TDI, smoking status, drinking status, ideal diet, sleep duration, physical activity, parental history of CMM, BMI, hypertension, dyslipidemia, and HbA1c. **Bold** indicates statistical significance. *Vascular diseases refer to CHD or stroke. 502 participants who were diagnosed with vascular diseases and T2DM on the same date were excluded in the analysis of sequential onset patterns of CMDs. HR, hazard ratio; OR, odds ratio; CI, confidence interval; SD, standard deviation, FCMD, first cardiometabolic disease; CMM, cardiometabolic multimorbidity; MVX, metabolic vulnerability index; SD, standard deviation; TDI, Townsend deprivation index; BMI, body mass index.

We further investigated the association of MVX with the sequential onset pattern of CMDs and uncovered a more prominent association in the pattern of T2DM followed by vascular diseases compared to the pattern of vascular diseases (CHD or stroke) followed by T2DM (**Figure 3**). Participants with highest sex-specific quartile of MVX suffered from a 59% higher risk (95%CI: 28%-97%) of developing T2DM followed by vascular diseases (**Additional File 1: Table S6**).

The restricted cubic spline models yielded similar results for these associations, suggesting the associations between higher MVX scores and higher risks of individual CMDs, CMM, and all-cause mortality (**Additional File 1: Figure S5**).

### 3.3 MVX and the transitions from healthy status to CMDs, CMM, and death

MSM analysis of transition pattern A further demonstrated a consistent positive association between MVX and the temporal progression of CMM (**Figure 4**). After full adjustment, each standard deviation increase of MVX score was associated with 9% (95%CI: 8%-10%), 5% (1%-10%), 11% (8%-14%), 10% (6%-13%), and 17% (8%-26%) higher risks of transitions from baseline to FCMD, from FCMD to CMM, from baseline to death, from FCMD to death, and from CMM to death, respectively (**Figure 4**). The associations of sex-specific quartiles of MVX, specific components of MVX, with the risk of transitions in pattern A are shown in **Additional File 1: Table S8-S9**. Restricted cubic spline analysis revealed similar results where higher MVX scores was associated with higher risks of baseline to FCMD, subsequently to CMM, and ultimately to death (**Additional File 1: Figure S6**). When taking CMD-specific associations into account (**Figure 4**), MVX was still significantly associated with the risk of most transitions in pattern B.

**Figure 4.**
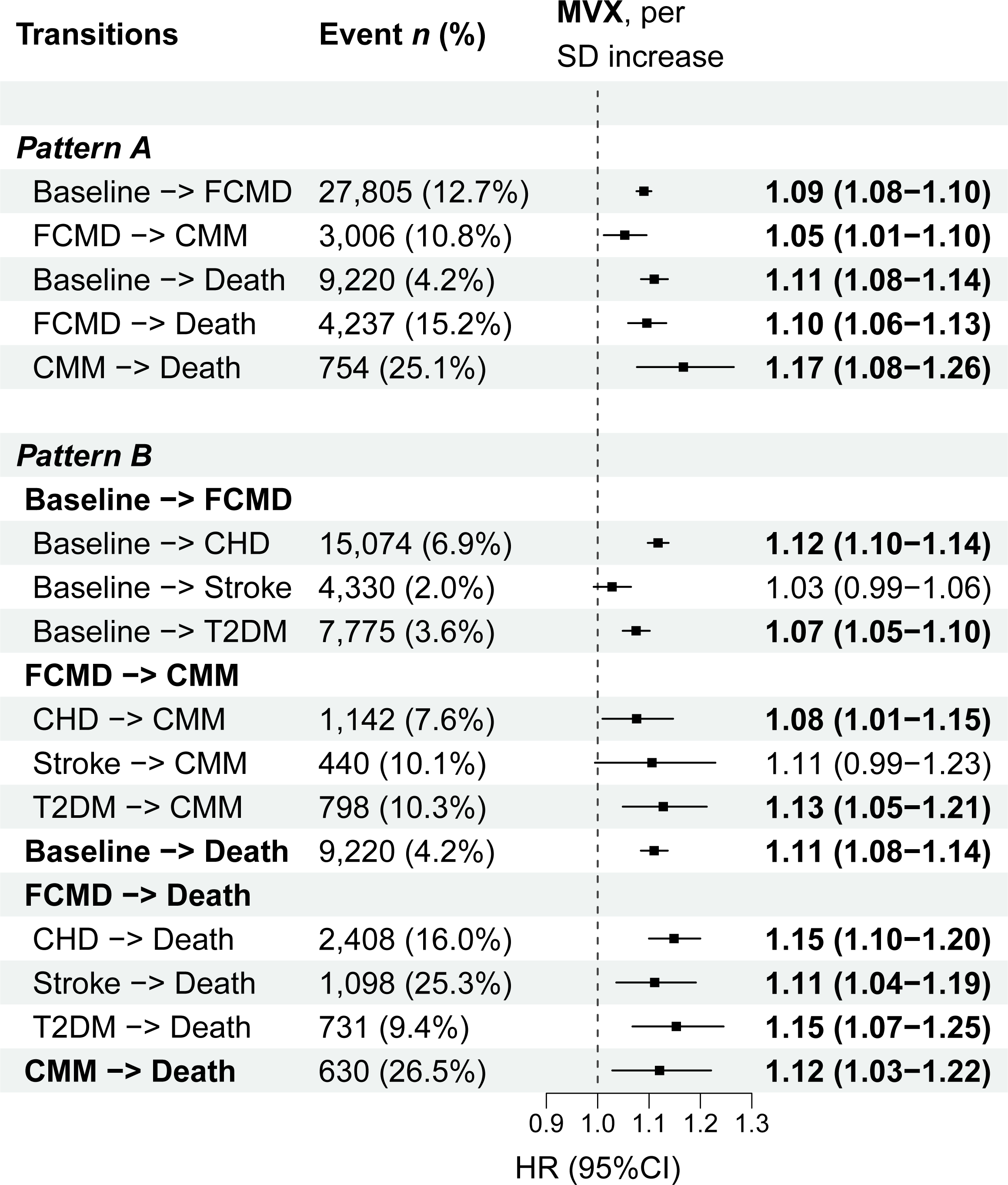
Hazard ratios (95%CIs) of MVX score for the risk of CMM progression in MSM analysis. The HRs (95%CIs) were adjusted for, adjusted for covariates as in Figure 3. **Bold** indicates statistical significance. HR, hazard ratio; CI, confidence interval; FCMD, first cardiometabolic disease; CMM, cardiometabolic multimorbidity; MVX, metabolic vulnerability index; SD, standard deviation; TDI, Townsend deprivation index; BMI, body mass index.

### 3.4 Stratified and sensitivity analyses

In stratified analyses, the associations between the MVX and incident CMDs, CMM, all-cause mortality, as well as the risks of each transition were similar across different subgroups, whereas potential effect modification of sex on the MVX-CMDs/CMM association was found (**Figure 5** and **Additional File 1: Figure S7**). The association between MVX and the outcomes was more pronounced in the female population.

**Figure 5.**
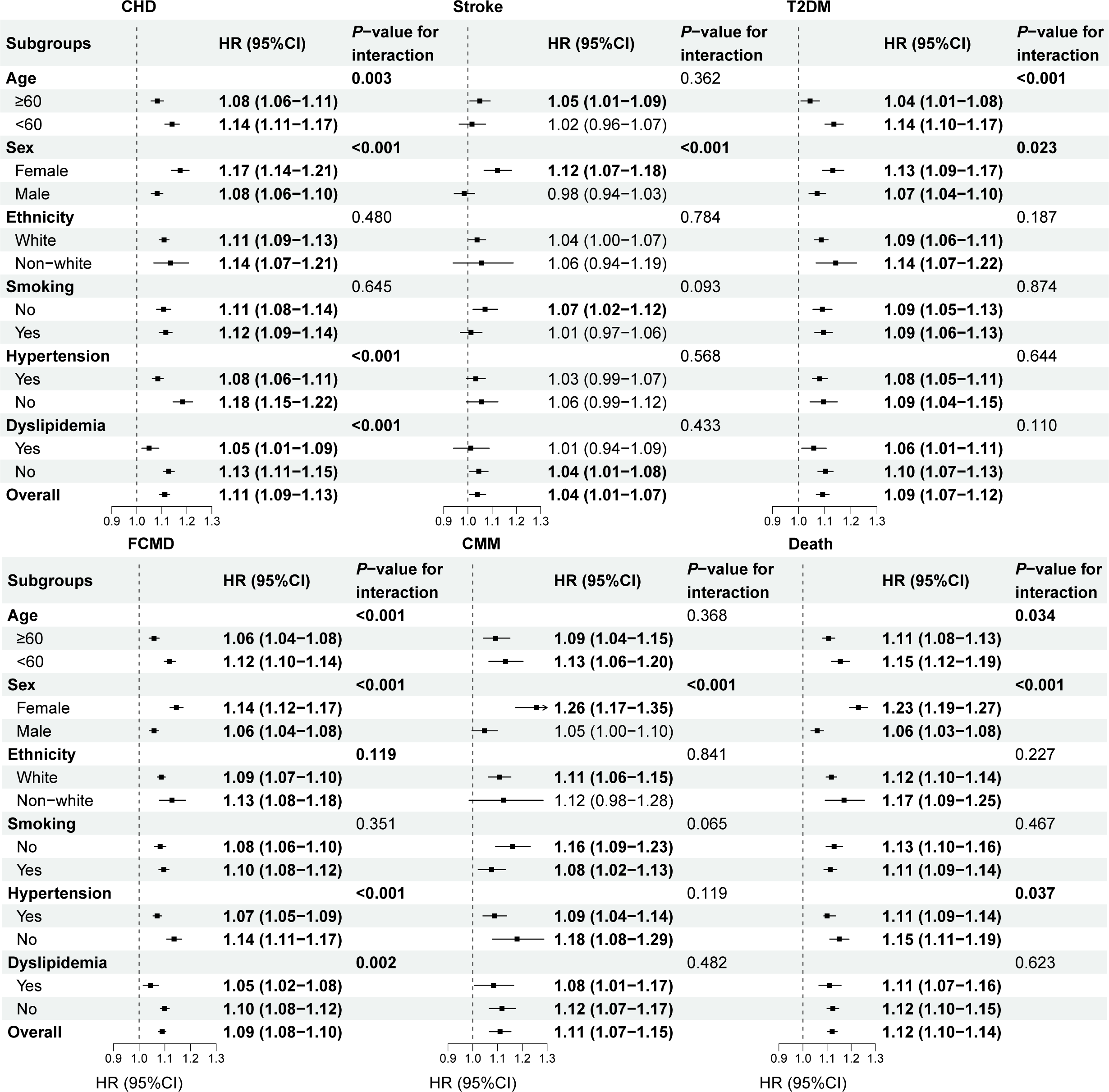
Hazard ratios (95% CIs) of MVX score for CMDs, CMM, and mortality in the stratified analysis. Results are HRs (95% CIs) per SD increase of MVX, adjusted for covariates as in Figure 3. **Bold** indicates statistical significance. CMD, cardiometabolic diseases; HR, hazard ratio; CI, confidence interval; SD, standard deviation; CHD, coronary heart disease; T2DM, type 2 diabetes mellitus; FCMD, first cardiometabolic disease; CMM, cardiometabolic multimorbidity; MVX, metabolic vulnerability index; SD, standard deviation.

We calculated the regression dilution ratio (RDR) to account for the fluctuation of MVX during follow-up. The RDR of MVX was 0.604 (95%CI: 0.590-0.618). When adjusted for the RDR, the associations of MVX with incident CMDs, their progression to CMM, and all-cause mortality became stronger, while maintaining statistical significance (**Additional File 1: Table S11**).

Results in the sensitivity analyses were similar to the main results (**Additional File 1: Table S12-S24**), which supported the robustness of study results.

## 4 Discussion

In the large-scale prospective cohort study, we elucidated the associations of MVX, a novel biomarker of systemic inflammation and metabolic malnutrition, with the risks of CMDs, CMM, and all-cause mortality based on a middle-aged and older UK population. The associations were more pronounced in females and in the sequential onset pattern of T2DM followed by vascular diseases. MSM analysis further illustrated consistent positive associations between MVX and temporal progression of CMM. Collectively, these results indicated the potential of MVX in the primary prevention and management of CMDs and CMM, thus satisfying the unmet need for precise assessment of inflammatory and metabolic status in the general population (**Figure 6**).

**Figure 6.**
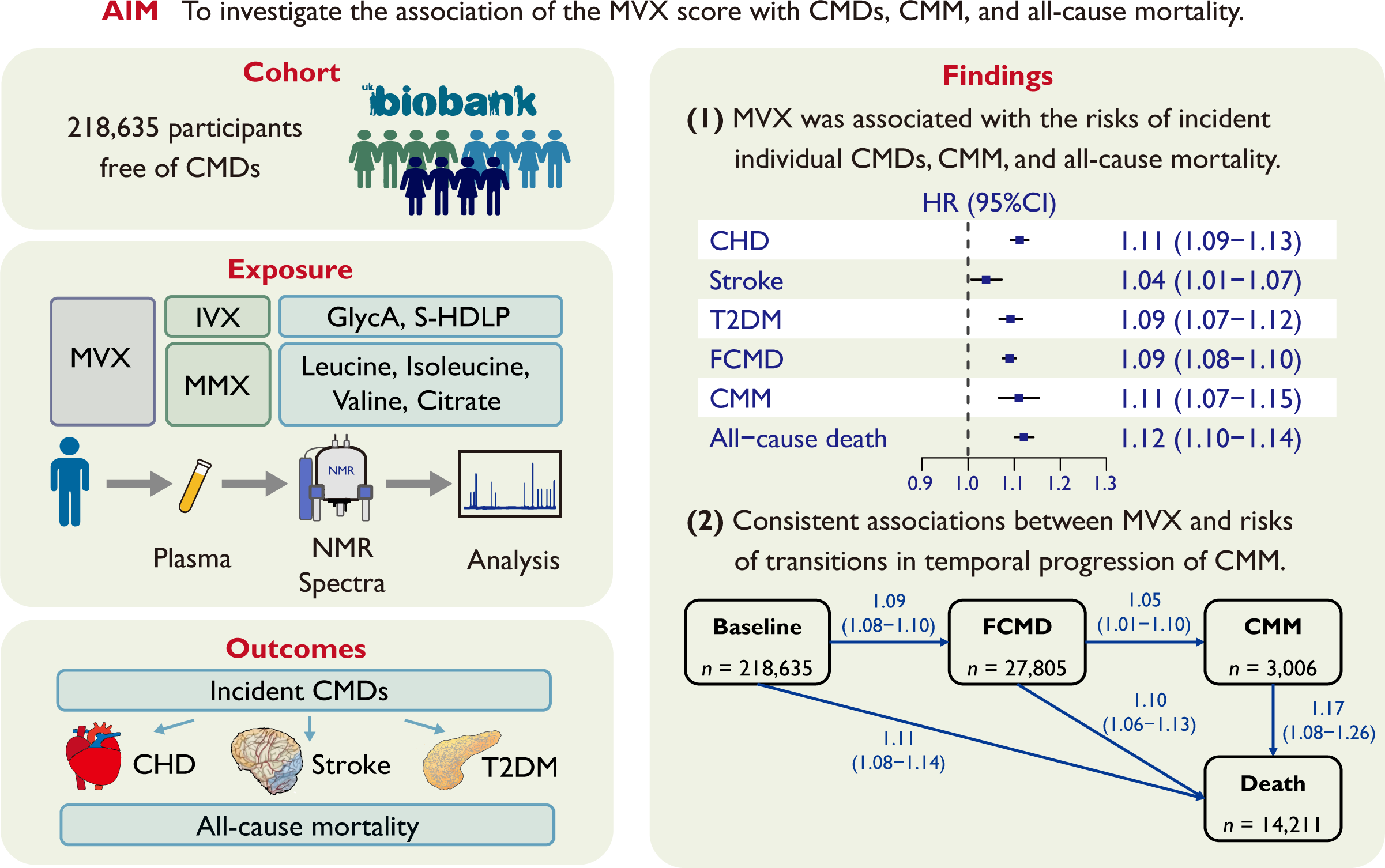
Associations of Metabolic Vulnerability Index with Cardiometabolic Diseases, Multimorbidity, and All-cause Mortality In a prospective cohort of 218,635 UK Biobank participants free of CMDs, MVX was calculated based on plasma metabolomics data. Each standard deviation increase of MVX was associated with 9% (95% confidence interval: 8%-10%), 11% (7%-15%), and 12% (10%-14%) higher risks of developing CMDs, CMM, and mortality, respectively. Multi-state model analysis further uncovered consistent associations between MVX and the risk of transitions from CMD-free to CMD, subsequently to CMM, and to death. CHD, coronary heart disease; CMD, cardiometabolic diseases; CMM, cardiometabolic multimorbidity; GlycA, glycoprotein acetyls; IVX, inflammatory vulnerability index; MMX, metabolic malnutrition index; MVX, metabolic vulnerability index; S-HDLP, small high-density lipoprotein particles; T2DM, type 2 diabetes mellitus.

The MVX score reflects the levels of systemic inflammation and metabolic malnutrition in a variety of chronic diseases. For instance, the mean MVX score was 41.3 in UKB participants, which was remarkably lower than that among populations with cardiovascular diseases, such as HF (mean MVX score: 63) [13] and CHD (mean MVX score: 50) [12]. Among UKB participants, those with higher MVX scores had higher prevalence of specific metabolic-related conditions, such as hypertension, dyslipidemia, and obesity, which was in line with previous literatures [12]. These results suggest that MVX could furnish an objective measurement of metabolic disease burden. In addition, our study indicated a positive correlation of MVX with insulin resistance, which gave mechanistic insight into the MVX-CMM association.

Both chronic low-grade sterile inflammation and metabolic malnutrition play critical roles in the pathogenesis and prognosis of CMDs [9, 34, 35]. For specific components in MVX, previous literature illustrated the significant associations of the two systemic inflammation markers with the incidence and prognosis of individual CMDs [20, 36–38]. GlycA originates from glycan residues of acute phase glycoproteins and reflects systemic inflammation [39], whereas S-HDLP exerts a majority of anti-inflammatory, anti-oxidant, and anti-thrombotic effects of HDL [40]. In terms of markers related to metabolic malnutrition, branched-chain amino acids (BCAAs), which are indicative of obesity-related metabolic disturbance, exhibit inverse correlations with cardiovascular disease and diabetes [41, 42]. Some studies also noticed citrate levels were associated with the risk of T2DM or cardiovascular mortality [43, 44].

Although several metabolomic and aforementioned biomarkers have been associated with CMDs and CMM, the predictive value of any single biomarker is quite limited. A composite metabolomic index could better refine risk stratification and management of CMDs. Given the complex association between inflammation and metabolic malnutrition in chronic diseases [45], it is inferable that a composite index of GlycA, S-HDLP, BCAAs, and citrate can serve as a promising predictor for the incidence, progression, and prognosis of CMDs and CMM. In a recent publication, Liu *et al.* uncovered that MVX independently predicts ASCVDs beyond traditional risk factors [46]. However, the prognostic value of MVX in the sequential progression of CMM remains to be investigated. By employing the MSM model, our study is able to assess heterogeneous MVX-CMM associations at different disease stages. Crucially, we revealed that higher MVX scores are not uniformly risky across all transitions, but are most prominently associated with the specific sequential pattern: from baseline healthy state, to T2DM, and then to vascular diseases, which is invisible in traditional or cross-sectional designs. This could be explained by the potential association between MVX and insulin resistance, coupled with the extended duration required to develop clinical macrovascular atherosclerosis [23]. From a clinical strategy perspective, these findings indicate that systemic inflammation and malnutrition could serve as potential targets for T2DM patients to halt the progression toward CHD or stroke. We also noticed a stronger MVX-CMD association among females in line with a recent study [14] More importantly, our study extended the generalizability of MVX, supporting its promising value in both primary and secondary prevention of individual CMDs and their multimorbidity.

Our study has several important strengths. Firstly, by leveraging a large-scale community-based prospective cohort with a long-term follow-up, our study uncovered the association of MVX with CMDs and CMM, which broadened the clinical applicability of MVX from secondary prevention of mortality in patients with cardiovascular diseases to primary prevention of CMDs in the general population. Secondly, most previous studies solely focused on the association of biomarker with the incidence or prognosis of a specific CMD [47]. By employing MSM which considers both various stages of natural CMM progression and competing risks [24], we investigated the transition-specific associations of MVX within a single analytic framework. The consistent associations across different transition stages further introduces MVX to a wider application spectrum. The prospective nature of the approach also reduced potential reverse causality and enhanced the comprehensiveness of our findings.

There were also several limitations to be noted when interpreting study results. First, our primary analysis was based on MVX assessed only at baseline, whereas MVX levels might fluctuate over time alongside lifestyle changes, medical treatments, or the progression of CMM itself. Although we performed regression dilution analysis to effectively account for the temporal changes of MVX during the follow-up, the clinical significance of its individual dynamic trajectories was not fully explored. Further studies with multiple serial measurement of MVX in a larger sample are needed. Second, for participants who entered multiple stages simultaneously, we calculated the prior state entrance date as the subsequent state entrance date minus 0.5 day in the primary analysis. Although several sensitivity analyses were performed and yielded similar results, the imprecision in the transition interval might result in biased estimation. Third, the observational nature of the study precluded causal inference, and we were unable to rule out residual confounding. Last, the UKB cohort mainly consisted of middle-aged and older White British, limiting the generalizability of our findings to other ethnicities.

## 5 Conclusion

Higher MVX scores were associated with increased risks of individual CMDs, their progressions to CMM, and eventually all-cause mortality. These results underscored the potential of MVX in the primary prevention and management of CMDs and CMM. Further investigation into the systemic inflammation and metabolic malnutrition is also urged for optimized management of CMDs and CMM.

## List of abbreviations

CHD: coronary heart disease
CMD: cardiometabolic disease
CMM: cardiometabolic multimorbidity
EDTA: ethylenediaminetetraacetic acid
GlycA: glycoprotein acetyls
HF: heart failure
IDI: integrated discrimination improvement
IVX: inflammation vulnerability index
MMX: metabolic malnutrition index
MSM: multi-state model
MVX: metabolic vulnerability index
NMR: nuclear magnetic resonance
NRI: net reclassification improvemen
S-HDLP: small high-density lipoprotein particles
T2DM: type 2 diabetes mellitus
UKB: UK Biobank

## Declarations

### Ethics approval and consent to participate

The UK Biobank study received ethical approval from the North West Multicenter Research Ethics Committee (reference number 11/NW/0382) and all participants have given written informed consents. This study has been approved by the UKB Access Management Team according to established access procedures (Application ID: 83421).

### Consent for publication

Not applicable.

### Availability of data and materials

All data analyzed in this study are publicly available in UK Biobank (https://www.ukbiobank.ac.uk/) after registration and proposal approval.

### Competing interests

The authors declare that they have no competing interests.

### Funding

This research is supported by the National Natural Science Foundation of China (grant 82370413 to YX), Noncommunicable Chronic Diseases-National Science and Technology Major Project (grant 2025ZD0547700 to MX), and Provincial and Ministry Joint Major Projects of National Health Commission of China (grant WKJ-ZJ-2405 to MX).

### Authors’ contributions

YX and MX contributed to the conception and design of this work. SZ, GY, HN, CZ, and YZ contributed to the analysis and interpretation of the data. YL and JS contributed to the visualization of the results. SZ and YL drafted the manuscript. GY, JS, YZ, XX, MX, and YX critically revised important intellectual content. YX and MX obtained funding for this work. XX and MX provided administrative support. All gave final approval and agreed to be accountable for all aspects of work ensuring integrity and accuracy.

## Acknowledgments

We sincerely thank all UKB staff and participants for their contribution.

## Additional File 1: Supplemental Materials

Additional File 1: Table S1-S24

Additional File 1: Figure S1-S7

Additional File 1: References #1-#2

## Notes

### Competing Interest Statement

The authors have declared no competing interest.

### Author Declarations

UK Biobank (https://www.ukbiobank.ac.uk/)

### Summary of Updates

This comprehensive revision involves adding new empirical evidence (figures/tables), optimizing existing data, and rewriting relevant sections.

