## Supplementary material for "Associations of Metabolic Vulnerability Index with Cardiometabolic Diseases, Multimorbidity, and All-cause Mortality": Tables S1-S24, Figures S1-S7

**Additional File 1:**

**Table S1.** Detailed definition of exclusion criteria.	6

**Table S2.** Source and detailed definitions of study outcomes.	11

**Table S3.** Source and detailed definitions of covariates.	13

**Table S4.** Testing results for the strict Markov assumption in multi-state models.	16

**Table S5.** Baseline characteristics stratified by sex-specific quartiles of MVX.	18

**Table S6.** Hazard ratios (95%CIs) of sex-specific quartiles of MVX for CMDs, CMM, and mortality.	25

**Table S7.** Hazard ratios (95%CIs) of individual MVX components for CMDs, CMM, and mortality.	28

**Table S8.** Discrimination ability of MVX for cardiometabolic diseases, multimorbidity, and all-cause mortality.	32

**Table S9.** Hazard ratios (95%CIs) of sex-specific quartiles of MVX for each transition in pattern A.	34

**Table S10.** Hazard ratios (95%CIs) of individual MVX components for each transition in pattern A.	35

**Table S11.** Hazard ratios (95%CIs) of MVX adjusted for regression dilution ratio.	37

**Table S12.** Hazard ratios (95%CIs) of MVX for each transition excluding participants who entered different states on the same date (*n* = 1,251).	39

**Table S13.** Hazard ratios (95%CIs) of MVX for each transition when calculating the entering date of the prior state using median transition interval time.	40

**Table S14.** Hazard ratios (95%CIs) of MVX for each transition after adding another transition from the baseline directly to CMM.	41

**Table S15.** Hazard ratios (95%CIs) of MVX for CMDs, CMM, mortality, and each transition after redefining T2DM as ICD-10 code E11 (specified T2DM).	42

**Table S16.** Hazard ratios (95%CIs) of MVX for CMDs, CMM, mortality, and each transition excluding participants with events occurring in the first two years of follow-up (*n* = 2,923).	44

**Table S17.** Hazard ratios (95%CIs) of MVX for CMDs, CMM, mortality, and each transition additionally adjusting for anti-hypertensive medication, cholesterol-lowering medication, albumin, and CRP.	46

**Table S18.** Hazard ratios (95%CIs) of MVX CMDs, CMM, mortality, and each transition excluding participants with missing values in covariates (*n* = 144,873).	48

**Table S19.** Hazard ratios (95%CIs) of MVX for CMDs, CMM, mortality, and each transition using data imputed with multiple imputation by chained equations.	50

**Table S20.** Hazard ratios (95%CIs) of MVX for CMDs, CMM, mortality, and each transition not excluding participants with cancer at baseline (*n* = 24,893).	52

**Table S21.** Hazard ratios (95%CIs) of MVX for each transition not excluding participants with CMDs at baseline (*n* = 29,893).	54

**Table S22.** Hazard ratios (95%CIs) of MVX for CMDs, CMM, mortality, and each transition additionally adjusted for statins use.	55

**Table S23.** Results of stratified analysis by the use of statins.	57

**Table S24.** Hazard ratios (95%CIs) of MVX for CMDs, CMM, mortality, and each transition additionally adjusted for serum phosphate.	59

**Figure S1.** *A priori* defined directed acyclic graph for confounders in the MVX-CMDs/CMM association.	61

**Figure S2.** Numbers and percentages of missing values in covariates.	63

**Figure S3.** Histogram of the MVX score in the UK Biobank cohort.	65

**Figure S4.** Results of the correlation analysis.	66

**Figure S5.** Restrictive cubic splines of MVX score for CMDs, CMM, and mortality.	68

**Figure S6.** Restrictive cubic splines of MVX score for each transition in pattern A.	70

**Figure S7.** Hazard ratios (95% CIs) of MVX score for each transition in the stratified analysis.	72

**References.**	74

**Table S1.** Detailed definition of exclusion criteria.

| **Variable** | **Definition / Description** | **Source (Field ID)** |
| --- | --- | --- |
| Coronary heart disease | No previous coronary heart disease (self-reported or previous diagnosis) | 6150 (self-reported)  131296 (I20 diagnosis date)  131298 (I21 diagnosis date)  131300 (I22 diagnosis date)  131302 (I23 diagnosis date)  131304 (I24 diagnosis date)  131306 (I25 diagnosis date) |
| Stroke | No previous stroke (self-reported or previous diagnosis) | 6150 (self-reported)  131360 (I60 diagnosis date)  131362 (I61 diagnosis date)  131364 (I62 diagnosis date)  131366 (I63 diagnosis date)  131368 (I64 diagnosis date)  131378 (I69 diagnosis date) |
| Diabetes | No previous diabetes (self-reported or previous diagnosis)  Baseline HbA1c<6.5% | 2443 (self-reported)  130706 (E10 diagnosis date)  130708 (E11 diagnosis date)  130710 (E12 diagnosis date)  130712 (E13 diagnosis date)  130714 (E14 diagnosis date) |
|  |  | 30750 (HbA1c) |
| Cancer | No previous cancer diagnosis | 40005 (cancer diagnosis date) |
| Study follow-up | Not withdrawn consent or lost follow-up | 190 (lost follow-up)  List of participants withdrawn consent (accessed Feb 23^rd^, 2024) |

**Table S2.** Source and detailed definitions of study outcomes.

| **Variable** | **Definition / Description** | **Source (Field ID)** |
| --- | --- | --- |
| All-cause mortality | Identified through linking to national death registries | 40000 (death date) |
| Type 2 diabetes | Ascertained from the “first occurrence of health outcomes” using linkage with death register, primary care, and hospital inpatient records.  ICD-10 code: both E11 (specified type 2 diabetes) and E14 (unspecified diabetes, which were primarily type 2 diabetes in UKB) in the primary analysis; only E11 in one sensitivity analysis | 130708 (E11 diagnosis date)  130714 (E14 diagnosis date) |
| Coronary heart disease | Ascertained from the “first occurrence of health outcomes” using linkage with death register, primary care, and hospital inpatient records.  ICD-10 code: I20-I25 (including myocardial infarction, angina, and ischemic cardiomyopathy) | 131296 (I20 diagnosis date)  131298 (I21 diagnosis date)  131300 (I22 diagnosis date)  131302 (I23 diagnosis date)  131304 (I24 diagnosis date)  131306 (I25 diagnosis date) |
| Stroke | Ascertained from the “first occurrence of health outcomes” using linkage with death register, primary care, and hospital inpatient records.  ICD-10 code: I60-I64 and I69 (including ischemic stroke, hemorrhagic stroke and *sequala* of stroke) | 131360 (I60 diagnosis date)  131362 (I61 diagnosis date)  131364 (I62 diagnosis date)  131366 (I63 diagnosis date)  131368 (I64 diagnosis date)  131378 (I69 diagnosis date) |

ICD-10, the International Statistical Classification of Diseases and Related Health Problems, 10^th^ Revision.

**Table S3.** Source and detailed definitions of covariates.

| **Variable** | **Definition / Description** | **Source (Field ID)** |
| --- | --- | --- |
| **Covariates adjusted in Cox regression and multi-state model analysis** | | |
| Age | Age at recruitment | 21022 |
| Sex |  | 31 |
| Ethnicity | White or others | 21000 |
| Education | College or university degree, or not | 6138 |
| Townsend deprivation index | Representing socio-economic status | 22189 |
| Smoking | Never, previous, or current | 20116 |
| Drinking | Never, previous, or current | 20117 |
| Ideal diet | Dietary score ≥4 points as proposed in the previous study^1^:   1) Vegetable intake at least four tablespoons each day 2) Fruit intake at least three pieces each day 3) Fish intake at least twice each week 4) Unprocessed red meat intake no more than twice each week 5) Processed meat intake no more than twice each week | 1289 (cooked vegetable)  1299 (raw vegetable)  1309 (fresh fruit)  1319 (dry fruit)  1329 (oily fish)  1339 (non-oily fish)  1349 (processed meat)  1369 (beef)  1379 (mutton)  1389 (pork) |
| Sleep duration | 7-8 hours/day as reference | 1160 |
| Physical activity above recommendation | 150 minutes of walking or moderate activity per week or 75 minutes of vigorous activity, or an equivalent combination | 22036 |
| Parental history of CMM | At least one parent suffering from at least two of heart diseases, stroke, or diabetes | 20107 (father)  20110 (mother) |
| Body mass index | Body mass (kg) divided by the square of height (m) | 21001 |
| Waist hip ratio | Waist circumference (cm) divided by hip circumference (cm) | 48 (waist circumference)  49 (hip circumference) |
| Hypertension | Defined as a self-reported history of hypertension, a previous diagnosis of hypertension (ICD-10 code: I10-I13 and I15), use of anti-hypertensive medication, or SBP ≥ 140mmHg, or DBP ≥ 90mmHg | 93 (SBP, manual reading)  94 (DBP, manual reading)  4079 (SBP, automated reading)  4080 (DBP, automated reading)  6150 (self-reported)  6177 (medication)  131286 (I10 diagnosis date)  131288 (I11 diagnosis date)  131290 (I12 diagnosis date)  131292 (I13 diagnosis date)  131294 (I15 diagnosis date) |
| Dyslipidemia | Defined as a previous diagnosis of dyslipidemia (ICD-10 code: E78), use of cholesterol-lowering medication | 6177 (medication)  130814 (E78 diagnosis date) |
| **Variables used in correlation analysis** | | |
| Albumin |  | 30600 |
| CRP |  | 30710 |
| HbA1c |  | 30750 |
| TyG index | TyG index = ln[triglycerides (mg/dL) * fasting glucose (mg/dL)]/2  Representing insulin resistance^2^ | 30740 (fasting glucose)  30870 (triglycerides) |
| Triglycerides |  | 30870 |
| Total cholesterol |  | 30690 |
| LDL-C |  | 30780 |
| HDL-C |  | 30760 |
| ApoA |  | 30630 |
| ApoB |  | 30640 |
| Lp(a) |  | 30790 |
| Serum phosphate |  |  |

BMI, body mass index; SBP, systolic blood pressure; DBP, diastolic blood pressure; CRP, C-reactive protein; HbA1c, glycated hemoglobulin A1c; LDL-C, low-density lipoprotein-cholesterol; HDL-C, high-density lipoprotein-cholesterol; Apo A, apolipoprotein A; Apo B, apolipoprotein B; Lp(a), lipoprotein (a); TyG index, triglyceride-glucose index.

**Table S4.** Testing results for the strict Markov assumption in multi-state models.

| **MSM Patterns** | **Transitions** | **GlycA** | **S-HDLP** | **IVX** | **Leucine** | **Isoleucine** | **Valine** | **Citrate** | **MMX** | **MVX** |
| --- | --- | --- | --- | --- | --- | --- | --- | --- | --- | --- |
| **Pattern A** | FCMD → CMM | *P*<0.001 | *P*<0.001 | *P*<0.001 | *P*<0.001 | *P*<0.001 | *P*<0.001 | *P*<0.001 | *P*<0.001 | *P*<0.001 |
|  | FCMD → Death | *P*<0.001 | *P*<0.001 | *P*<0.001 | *P*<0.001 | *P*<0.001 | *P*<0.001 | *P*<0.001 | *P*<0.001 | *P*<0.001 |
|  | CMM → Death | *P*<0.001 | *P*<0.001 | *P*<0.001 | *P*<0.001 | *P*<0.001 | *P*<0.001 | *P*<0.001 | *P*<0.001 | *P*<0.001 |
| **Pattern B** | CHD → CMM | *P*<0.001 | *P*<0.001 | *P*<0.001 | *P*<0.001 | *P*<0.001 | *P*<0.001 | *P*<0.001 | *P*<0.001 | *P*<0.001 |
|  | CHD → Death | *P*<0.001 | *P*<0.001 | *P*<0.001 | *P*<0.001 | *P*<0.001 | *P*<0.001 | *P*<0.001 | *P*<0.001 | *P*<0.001 |
|  | Stroke → CMM | *P*<0.001 | *P*<0.001 | *P*<0.001 | *P*<0.001 | *P*<0.001 | *P*<0.001 | *P*<0.001 | *P*<0.001 | *P*<0.001 |
|  | Stroke → Death | *P*<0.001 | *P*<0.001 | *P*<0.001 | *P*<0.001 | *P*<0.001 | *P*<0.001 | *P*<0.001 | *P*<0.001 | *P*<0.001 |
|  | T2DM → CMM | *P*<0.001 | *P*<0.001 | *P*<0.001 | *P*<0.001 | *P*<0.001 | *P*<0.001 | *P*<0.001 | *P*<0.001 | *P*<0.001 |
|  | T2DM → Death | *P*<0.001 | *P*<0.001 | *P*<0.001 | *P*<0.001 | *P*<0.001 | *P*<0.001 | *P*<0.001 | *P*<0.001 | *P*<0.001 |
|  | CMM → Death | *P*<0.001 | *P*<0.001 | *P*<0.001 | *P*<0.001 | *P*<0.001 | *P*<0.001 | *P*<0.001 | *P*<0.001 | *P*<0.001 |

Results are *P*-values for "time since entry into the current state" (duration of the previous conditions) term in the fully-adjusted transition regression models. These data demonstrated that the "time since entry into the current state" was significantly associated with the risk of subsequent transitions across our models, directly rejecting the strict Markov assumption and strongly justifying the use of the semi-Markov approach.

**Table S5.** Baseline characteristics stratified by sex-specific quartiles of MVX.

| **Characteristic** | **Overall,** *n* = 218,635 | **Q1,** *n* = 54,659 | **Q2,** *n* = 54,659 | **Q3,** *n* = 54,658 | **Q4,** *n* = 54,659 | ***P*-value** |
| --- | --- | --- | --- | --- | --- | --- |
| **Sociodemographic variables** |  |  |  |  |  |  |
| **Age**, year | 55.75 (8.09) | 54.69 (8.12) | 55.75 (8.10) | 56.15 (8.07) | 56.41 (7.96) | **<0.001** |
| **Female**, *n* (%) | 120,483 (55.1%) | 30,121 (55.1%) | 30,121 (55.1%) | 30,120 (55.1%) | 30,121 (55.1%) | >0.999 |
| **Ethnicity**, *n* (%) |  |  |  |  |  | **<0.001** |
| White | 199,098 (91.5%) | 48,908 (89.9%) | 49,772 (91.4%) | 50,069 (92.0%) | 50,349 (92.5%) |  |
| Non-white | 18,597 (8.54%) | 5,498 (10.1%) | 4,659 (8.56%) | 4,366 (8.02%) | 4,074 (7.49%) |  |
| **Education level**, *n* (%) |  |  |  |  |  | **<0.001** |
| College or university degree | 72,751 (33.6%) | 22,603 (41.7%) | 18,793 (34.7%) | 16,931 (31.3%) | 14,424 (26.7%) |  |
| Less than college or university | 143,540 (66.4%) | 31,549 (58.3%) | 35,321 (65.3%) | 37,116 (68.7%) | 39,554 (73.3%) |  |
| **Townsend deprivation index** | -2.27 (-3.71, 0.27) | -2.38 (-3.79, 0.00) | -2.36 (-3.76, 0.07) | -2.26 (-3.71, 0.27) | -2.04 (-3.58, 0.77) | **<0.001** |
| **Lifestyle factors** |  |  |  |  |  |  |
| **Smoking status**, *n* (%) |  |  |  |  |  | **<0.001** |
| Never | 123,224 (56.6%) | 32,965 (60.6%) | 31,620 (58.1%) | 30,429 (55.9%) | 28,210 (51.9%) |  |
| Previous | 71,946 (33.0%) | 17,511 (32.2%) | 17,918 (32.9%) | 18,027 (33.1%) | 18,490 (34.0%) |  |
| Current | 22,530 (10.3%) | 3,958 (7.27%) | 4,903 (9.01%) | 5,973 (11.0%) | 7,696 (14.1%) |  |
| **Drinking status**, *n* (%) |  |  |  |  |  | **<0.001** |
| Never | 8,629 (3.96%) | 1,615 (2.96%) | 2,092 (3.83%) | 2,281 (4.18%) | 2,641 (4.84%) |  |
| Previous | 6,815 (3.12%) | 1,374 (2.52%) | 1,642 (3.01%) | 1,727 (3.17%) | 2,072 (3.80%) |  |
| Current | 202,718 (92.9%) | 51,559 (94.5%) | 50,822 (93.2%) | 50,536 (92.7%) | 49,801 (91.4%) |  |
| **Ideal diet**, *n* (%) | 87,674 (41.8%) | 23,487 (44.4%) | 22,449 (42.7%) | 21,684 (41.4%) | 20,054 (38.5%) | **<0.001** |
| **Sleep duration**, *n* (%) |  |  |  |  |  | **<0.001** |
| ≤6h | 52,690 (24.3%) | 12,475 (22.9%) | 12,939 (23.8%) | 13,198 (24.3%) | 14,078 (26.0%) |  |
| 7-8h | 149,413 (68.8%) | 38,850 (71.4%) | 37,901 (69.7%) | 37,186 (68.5%) | 35,476 (65.5%) |  |
| ≥9h | 15,095 (6.95%) | 3,073 (5.65%) | 3,510 (6.46%) | 3,900 (7.18%) | 4,612 (8.51%) |  |
| **Physical activity above recommendation**, *n* (%) | 140,299 (82.3%) | 37,857 (85.1%) | 35,670 (82.8%) | 34,272 (81.5%) | 32,500 (79.4%) | **<0.001** |
| **Medical conditions** |  |  |  |  |  |  |
| **Hypertension**, *n* (%) | 113,776 (52.2%) | 22,833 (41.9%) | 27,335 (50.2%) | 29,782 (54.7%) | 33,826 (62.1%) | **<0.001** |
| **Dyslipidemia**, *n* (%) | 26,198 (12.0%) | 5,220 (9.59%) | 6,215 (11.4%) | 6,779 (12.5%) | 7,984 (14.7%) | **<0.001** |
| **Anti-hypertensive medication**, *n* (%) | 16,424 (7.55%) | 3,167 (5.82%) | 4,021 (7.39%) | 4,345 (7.99%) | 4,891 (8.99%) | **<0.001** |
| **Cholesterol-lowering medication**, *n* (%) | 12,702 (5.84%) | 2,688 (4.94%) | 3,112 (5.72%) | 3,277 (6.02%) | 3,625 (6.66%) | **<0.001** |
| **Parental history of CMM**, *n* (%) | 27,124 (13.6%) | 6,335 (12.6%) | 6,732 (13.5%) | 7,023 (14.1%) | 7,034 (14.2%) | **<0.001** |
| **Physical examinations** |  |  |  |  |  |  |
| **BMI**, kg/m^2^ | 27.11 (4.55) | 25.79 (3.91) | 26.77 (4.30) | 27.44 (4.56) | 28.45 (4.95) | **<0.001** |
| **Waist-hip ratio** | 0.87 (0.09) | 0.85 (0.08) | 0.86 (0.09) | 0.87 (0.09) | 0.89 (0.09) | **<0.001** |
| **SBP**, mmHg | 137.49 (18.59) | 133.72 (18.03) | 136.65 (18.27) | 138.53 (18.54) | 141.07 (18.72) | **<0.001** |
| **DBP**, mmHg | 82.38 (10.12) | 80.24 (9.97) | 81.89 (9.96) | 82.92 (10.01) | 84.46 (10.07) | **<0.001** |
| **Laboratory Tests** |  |  |  |  |  |  |
| **Albumin**, g/L | 45.26 (2.59) | 45.41 (2.52) | 45.27 (2.54) | 45.23 (2.58) | 45.12 (2.72) | **<0.001** |
| **CRP**, mg/L | 1.28 (0.64, 2.63) | 0.76 (0.41, 1.42) | 1.11 (0.59, 2.13) | 1.45 (0.76, 2.84) | 2.29 (1.15, 4.66) | **<0.001** |
| **Fasting glucose**, mmol/L | 4.95 (0.66) | 4.90 (0.64) | 4.94 (0.63) | 4.96 (0.65) | 5.02 (0.72) | **<0.001** |
| **HbA1c**, % | 5.34 (0.34) | 5.25 (0.33) | 5.32 (0.33) | 5.36 (0.33) | 5.43 (0.34) | **<0.001** |
| **Total cholesterol**, mmol/L | 5.81 (1.08) | 5.56 (1.02) | 5.70 (1.03) | 5.85 (1.05) | 6.13 (1.14) | **<0.001** |
| **LDL-C**, mmol/L | 3.64 (0.83) | 3.43 (0.77) | 3.57 (0.79) | 3.69 (0.81) | 3.88 (0.87) | **<0.001** |
| **HDL-C**, mmol/L | 1.47 (0.38) | 1.56 (0.39) | 1.48 (0.38) | 1.44 (0.37) | 1.39 (0.34) | **<0.001** |
| **Triglycerides**, mmol/L | 1.45 (1.03, 2.10) | 1.13 (0.86, 1.52) | 1.35 (1.00, 1.84) | 1.57 (1.12, 2.19) | 2.02 (1.39, 2.88) | **<0.001** |
| **Apo A**, g/L | 1.55 (0.27) | 1.59 (0.28) | 1.55 (0.27) | 1.53 (0.26) | 1.52 (0.25) | **<0.001** |
| **Apo B**, g/L | 1.05 (0.23) | 0.98 (0.21) | 1.03 (0.22) | 1.06 (0.23) | 1.13 (0.25) | **<0.001** |
| **Lp(a)**, nmol/L | 20.88 (9.60, 61.22) | 21.10 (9.40, 67.76) | 21.20 (9.70, 63.66) | 21.00 (9.73, 59.60) | 20.20 (9.50, 55.60) | **<0.001** |
| **TyG index** | 4.68 (0.27) | 4.55 (0.23) | 4.64 (0.24) | 4.71 (0.25) | 4.83 (0.28) | **<0.001** |
| **Components of MVX** |  |  |  |  |  |  |
| **GlycA**, μmol/L | 806.42 (118.34) | 698.56 (64.88) | 767.74 (65.65) | 824.42 (76.45) | 934.95 (108.09) | **<0.001** |
| **S-HDLP**, μmol/L | 9.82 (1.32) | 9.52 (1.27) | 9.61 (1.19) | 9.81 (1.20) | 10.35 (1.47) | **<0.001** |
| **IVX score** | 41.19 (18.47) | 27.25 (13.06) | 35.54 (13.62) | 42.87 (13.57) | 59.11 (16.57) | **<0.001** |
| **Isoleucine**, μmol/L | 50.63 (18.00) | 50.72 (15.04) | 49.70 (16.54) | 49.75 (18.35) | 52.37 (21.34) | **<0.001** |
| **Leucine**, μmol/L | 103.33 (28.48) | 106.68 (22.47) | 102.09 (25.65) | 100.78 (29.25) | 103.77 (34.75) | **<0.001** |
| **Valine**, μmol/L | 208.56 (42.77) | 209.46 (32.54) | 205.04 (37.80) | 205.14 (43.82) | 214.59 (53.36) | **<0.001** |
| **Citrate,** μmol/L | 65.09 (12.92) | 63.18 (12.79) | 64.83 (12.69) | 65.48 (12.76) | 66.89 (13.16) | **<0.001** |
| **MMX score** | 45.85 (16.00) | 39.68 (14.06) | 45.14 (14.65) | 48.57 (15.76) | 49.99 (17.32) | **<0.001** |

Results are number (percentage%) for categorical variables and mean (standard deviation, SD) or median (interquartile range, IQR) for continuous variables. TyG index (reflexing insulin resistance) was calculated as ln[triglycerides (mg/dL) × fasting glucose (mg/dL)]. CMM, cardiometabolic multimorbidity; BMI, body mass index; SBP, systolic blood pressure; DBP, diastolic blood pressure; CRP, C-reactive protein; HbA1c, glycated hemoglobulin A1c; LDL-C, low-density lipoprotein-cholesterol; HDL-C, high-density lipoprotein-cholesterol; Apo A, apolipoprotein A; Apo B, apolipoprotein B; Lp(a), lipoprotein (a); TyG index, triglyceride-glucose index; GlycA, glycoprotein acetyls; S-HDLP, small high-density lipoprotein particles; IVX, inflammation vulnerability index; MMX, metabolic malnutrition index; MVX, metabolic vulnerability index.

**Table S6.** Hazard ratios (95%CIs) of sex-specific quartiles of MVX for CMDs, CMM, and mortality.

|  | **Sex-specific quartiles of MVX** | | | |
| --- | --- | --- | --- | --- |
|  | **Q1** | **Q2** | **Q3** | **Q4** |
| **Individual CMDs** |  |  |  |  |
| CHD | 1.00 (Reference) | **1.10 (1.05-1.16)** | **1.17 (1.12-1.22)** | **1.30 (1.24-1.36)** |
| Stroke | 1.00 (Reference) | 1.04 (0.95-1.13) | 1.05 (0.97-1.14) | **1.10 (1.02-1.20)** |
| T2DM | 1.00 (Reference) | **1.09 (1.01-1.17)** | **1.11 (1.03-1.19)** | **1.23 (1.15-1.31)** |
| **CMM status** |  |  |  |  |
| FCMD | 1.00 (Reference) | **1.06 (1.03-1.11)** | **1.10 (1.06-1.14)** | **1.22 (1.17-1.26)** |
| CMM | 1.00 (Reference) | **1.16 (1.03-1.32)** | **1.20 (1.06-1.35)** | **1.35 (1.20-1.51)** |
| **All-cause death** | 1.00 (Reference) | **1.11 (1.05-1.17)** | **1.17 (1.11-1.23)** | **1.32 (1.26-1.39)** |
| Death without CMDs | 1.00 (Reference) | **1.11 (1.04-1.18)** | **1.17 (1.10-1.24)** | **1.29 (1.22-1.38)** |
| Death with single CMD | 1.00 (Reference) | **1.13 (1.02-1.24)** | **1.19 (1.08-1.31)** | **1.40 (1.27-1.53)** |
| Death with CMM | 1.00 (Reference) | 1.12 (0.87-1.43) | 1.16 (0.92-1.48) | **1.39 (1.10-1.74)** |
| **Sequential onset pattern of CMDs*** |  |  |  |  |
| T2DM only | 1.00 (Reference) | 1.06 (0.97-1.15) | 1.08 (0.99-1.17) | **1.20 (1.10-1.30)** |
| Vascular diseases only | 1.00 (Reference) | **1.08 (1.03-1.13)** | **1.14 (1.09-1.20)** | **1.27 (1.21-1.33)** |
| T2DM followed by vascular diseases | 1.00 (Reference) | 1.11 (0.87-1.41) | **1.38 (1.10-1.72)** | **1.59 (1.28-1.97)** |
| Vascular diseases followed by T2DM | 1.00 (Reference) | 1.20 (0.94-1.53) | 1.05 (0.82-1.33) | 1.23 (0.98-1.55) |

Results are HRs (95%CIs) for the MVX-CMDs/CMM/mortality associations and ORs (95%CIs) for the MVX-sequential onset pattern associations, adjusted for age, sex, ethnicity, education level, TDI, smoking status, drinking status, diet, sleep duration, physical activity, parental history of CMM, hypertension, dyslipidemia, BMI, and HbA1c. **Bold** indicates statistical significance. *502 participants who were diagnosed with vascular diseases and T2DM on the same date were excluded in the analysis. HR, hazard ratio; OR, odds ratio; CI, confidence interval; SD, standard deviation; FCMD, first cardiometabolic disease; CHD, coronary heart disease; T2DM, type 2 diabetes mellitus; CMM, cardiometabolic multimorbidity; GlycA, glycoprotein acetyls; S-HDLP, small high-density lipoprotein particles; TDI, Townsend deprivation index; BMI, body mass index.

**Table S7.** Hazard ratios (95%CIs) of individual MVX components for CMDs, CMM, and mortality.

|  | **GlycA** | **S-HDLP** | **IVX** | **Leucine** | **Isoleucine** | **Valine** | **Citrate** | **MMX** |
| --- | --- | --- | --- | --- | --- | --- | --- | --- |
| **Individual CMDs** |  |  |  |  |  |  |  |  |
| CHD | **1.13 (1.12-1.15)** | **0.97 (0.95-0.98)** | **1.14 (1.12-1.16)** | **1.02 (1.01-1.04)** | **1.03 (1.02-1.05)** | **1.02 (1.01-1.04)** | 0.99 (0.97-1.00) | 0.99 (0.97-1.01) |
| Stroke | **1.03 (1.00-1.07)** | **0.92 (0.90-0.95)** | 1.02 (0.99-1.06) | 0.99 (0.96-1.02) | 1.01 (0.98-1.04) | 0.98 (0.95-1.01) | 0.99 (0.96-1.02) | **1.06 (1.02-1.09)** |
| T2DM | **1.14 (1.12-1.16)** | 0.98 (0.96-1.00) | **1.15 (1.13-1.18)** | **1.12 (1.09-1.14)** | **1.11 (1.09-1.13)** | **1.17 (1.15-1.2)** | 0.99 (0.97-1.01) | **0.89 (0.86-0.91)** |
| **CMM status** |  |  |  |  |  |  |  |  |
| FCMD | **1.12 (1.10-1.13)** | **0.96 (0.95-0.97)** | **1.12 (1.10-1.13)** | **1.04 (1.03-1.06)** | **1.05 (1.04-1.06)** | **1.06 (1.05-1.07)** | 0.99 (0.98-1.00) | **0.98 (0.96-0.99)** |
| CMM | **1.12 (1.08-1.16)** | **0.91 (0.88-0.95)** | **1.13 (1.08-1.18)** | **1.04 (1.01-1.08)** | **1.07 (1.03-1.10)** | **1.08 (1.04-1.11)** | 0.99 (0.96-1.03) | 1.01 (0.96-1.06) |
| **All-cause death** | **1.10 (1.08-1.12)** | **0.88 (0.87-0.90)** | 1.07 (1.05-1.10) | **0.91 (0.89-0.93)** | **0.95 (0.94-0.97)** | **0.89 (0.87-0.91)** | **1.02 (1.00-1.04)** | **1.15 (1.13-1.18)** |
| Death without CMDs | **1.08 (1.06-1.1)** | **0.89 (0.87-0.91)** | 1.05 (1.03-1.08) | **0.90 (0.88-0.92)** | **0.94 (0.92-0.96)** | **0.88 (0.86-0.9)** | 1.02 (1.00-1.04) | **1.15 (1.12-1.17)** |
| Death with single CMD | **1.13 (1.10-1.17)** | **0.89 (0.87-0.92)** | 1.12 (1.08-1.17) | **0.91 (0.88-0.94)** | **0.95 (0.92-0.98)** | **0.90 (0.87-0.93)** | 1.02 (0.99-1.05) | **1.15 (1.10-1.19)** |
| Death with CMM | **1.13 (1.05-1.22)** | **0.83 (0.77-0.89)** | 1.13 (1.03-1.23) | 1.01 (0.94-1.09) | 1.04 (0.97-1.12) | 1.02 (0.95-1.10) | 1.00 (0.93-1.08) | **1.20 (1.10-1.31)** |
| **Sequential onset pattern of CMDs*** |  |  |  |  |  |  |  |  |
| T2DM only | **1.16 (1.13-1.19)** | 0.99 (0.96-1.01) | **1.17 (1.13-1.20)** | **1.15 (1.12-1.18)** | **1.14 (1.11-1.17)** | **1.22 (1.19-1.25)** | 1.00 (0.97-1.02) | **0.87 (0.84-0.90)** |
| Vascular diseases only | **1.13 (1.11-1.15)** | **0.97 (0.95-0.98)** | **1.13 (1.11-1.15)** | **1.03 (1.01-1.04)** | **1.04 (1.02-1.06)** | **1.02 (1.01-1.04)** | 0.98 (0.97-1.00) | 1.00 (0.98-1.02) |
| T2DM followed by vascular diseases | **1.24 (1.16-1.32)** | **0.92 (0.86-0.98)** | **1.24 (1.15-1.35)** | **1.09 (1.02-1.17)** | **1.12 (1.05-1.19)** | **1.17 (1.10-1.25)** | 0.99 (0.92-1.06) | 0.96 (0.87-1.05) |
| Vascular diseases followed by T2DM | **1.15 (1.07-1.24)** | **0.90 (0.83-0.97)** | **1.16 (1.06-1.26)** | **1.11 (1.04-1.19)** | **1.12 (1.05-1.20)** | **1.17 (1.09-1.25)** | 0.97 (0.91-1.05) | 0.95 (0.86-1.05) |

Results are HRs (95%CIs) for the MVX-CMDs/CMM/mortality associations and ORs (95%CIs) for the MVX-sequential onset pattern associations, per one SD increase of each MVX component, adjusted for age, sex, ethnicity, education level, TDI, smoking status, drinking status, diet, sleep duration, physical activity, parental history of CMM, hypertension, dyslipidemia, BMI, and HbA1c. **Bold** indicates statistical significance. *502 participants who were diagnosed with vascular diseases and T2DM on the same date were excluded in the analysis. HR, hazard ratio; OR, odds ratio; CI, confidence interval; SD, standard deviation; GlycA, glycoprotein acetyls; S-HDLP, small high-density lipoprotein particles; IVX, inflammation vulnerability index; MMX, metabolic malnutrition index; FCMD, first cardiometabolic disease; CHD, coronary heart disease; T2DM, type 2 diabetes mellitus; CMM, cardiometabolic multimorbidity; TDI, Townsend deprivation index; BMI, body mass index.

**Table S8.** Discrimination ability of MVX for cardiometabolic diseases, multimorbidity, and all-cause mortality.

| **Outcomes** | **Uno’s C-index** | | **Integrated discrimination improvement** | | **Net reclassification improvement** | |
| --- | --- | --- | --- | --- | --- | --- |
|  | Model without MVX | Model with MVX | Point estimate (95% CI) | *P*-value | Point estimate (95% CI) | *P*-value |
| **CHD** | 0.707 (0.698~0.716) | 0.709 (0.699~0.718) | 0.036% (0.017% ~ 0.055%) | **<0.001** | 3.89% (2.64% ~ 4.73%) | **<0.001** |
| **Stroke** | 0.713 (0.706~0.720) | 0.713 (0.706~0.720) | 0.000% (-0.002% ~ 0.008%) | >0.999 | -0.23% (-2.23% ~ 1.63%) | 0.970 |
| **T2DM** | 0.838 (0.834~0.843) | 0.839 (0.834~0.844) | 0.018% (-0.007% ~ 0.053%) | 0.198 | 4.64% (2.61% ~ 6.25%) | **<0.001** |
| **FCMD** | 0.723 (0.718~0.728) | 0.724 (0.719~0.729) | 0.029% (0.008% ~ 0.052%) | **<0.001** | 3.20% (2.30% ~ 3.96%) | **<0.001** |
| **CMM** | 0.815 (0.808~0.823) | 0.817 (0.809~0.824) | 0.006% (-0.013% ~ 0.029%) | 0.495 | 4.03% (0.71% ~ 6.46%) | **0.020** |
| **All-cause mortality** | 0.742 (0.738~0.746) | 0.744 (0.740~0.748) | 0.042% (0.023% ~ 0.067%) | <0.001 | 4.45% (2.94% ~ 5.62%) | **<0.001** |

Model without MVX includes age at baseline, sex, ethnicity, educational level, Townsend deprivation index, smoking status, drinking status, diet, sleep duration, physical activity, body mass index, parental history of CMM, history of hypertension and dyslipidemia, and hemoglobin A1c levels. **Bold** indicates statistical significance. CMD, cardiometabolic diseases; CHD, coronary heart disease; T2DM, type 2 diabetes mellitus; FCMD, first cardiometabolic disease; CMM, cardiometabolic multimorbidity; CI, confidence interval; MVX, metabolic vulnerability index.

**Table S9.** Hazard ratios (95%CIs) of sex-specific quartiles of MVX for each transition in pattern A.

|  | **Sex-specific quartiles of MVX** | | | |
| --- | --- | --- | --- | --- |
|  | **Q1** | **Q2** | **Q3** | **Q4** |
| Baseline → FCMD | 1.00 (Reference) | **1.05 (1.01-1.09)** | **1.10 (1.06-1.14)** | **1.23 (1.19-1.28)** |
| FCMD → CMM | 1.00 (Reference) | **1.13 (1.00-1.28)** | 1.12 (0.99-1.26) | **1.20 (1.06-1.34)** |
| Baseline → Death | 1.00 (Reference) | **1.11 (1.05-1.19)** | **1.17 (1.10-1.25)** | **1.29 (1.21-1.37)** |
| FCMD → Death | 1.00 (Reference) | **1.15 (1.04-1.27)** | 1.09 (0.99-1.21) | **1.21 (1.10-1.33)** |
| CMM → Death | 1.00 (Reference) | **1.31 (1.01-1.69)** | 1.23 (0.96-1.59) | **1.32 (1.04-1.68)** |

Results are HRs (95%CIs), adjusted for age, sex, ethnicity, education level, TDI, smoking status, drinking status, diet, sleep duration, physical activity, parental history of CMM, hypertension, dyslipidemia, BMI, and HbA1c. **Bold** indicates statistical significance. HR, hazard ratio; CI, confidence interval; SD, standard deviation; FCMD, first cardiometabolic disease; CMM, cardiometabolic multimorbidity; GlycA, glycoprotein acetyls; S-HDLP, small high-density lipoprotein particles; TDI, Townsend deprivation index; BMI, body mass index.

**Table S10.** Hazard ratios (95%CIs) of individual MVX components for each transition in pattern A.

| **Transitions** | **GlycA** | **S-HDLP** | **IVX** | **Leucine** | **Isoleucine** | **Valine** | **Citrate** | **MMX** |
| --- | --- | --- | --- | --- | --- | --- | --- | --- |
| Baseline → FCMD | **1.10 (1.09-1.11)** | **0.93 (0.92-0.94)** | **1.13 (1.11-1.14)** | **1.03 (1.02-1.04)** | **1.04 (1.03-1.05)** | **1.04 (1.03-1.05)** | **0.97 (0.96-0.99)** | **0.95 (0.94-0.97)** |
| FCMD → CMM | **1.06 (1.02-1.10)** | 0.99 (0.95-1.02) | 1.04 (0.99-1.09) | 1.02 (0.99-1.06) | 1.03 (1.00-1.07) | **1.04 (1.00-1.07)** | 1.03 (0.99-1.06) | **1.09 (1.04-1.14)** |
| Baseline → Death | **1.10 (1.07-1.12)** | **0.90 (0.88-0.92)** | **1.05 (1.03-1.08)** | **0.92 (0.90-0.94)** | **0.95 (0.93-0.97)** | **0.90 (0.88-0.92)** | **1.03 (1.01-1.05)** | **1.16 (1.13-1.18)** |
| FCMD → Death | **1.12 (1.08-1.15)** | **1.06 (1.03-1.09)** | 1.02 (0.98-1.06) | **0.94 (0.91-0.97)** | **0.95 (0.92-0.98)** | **0.92 (0.89-0.95)** | **1.10 (1.07-1.13)** | **1.30 (1.25-1.34)** |
| CMM → Death | **1.16 (1.08-1.25)** | 1.06 (0.99-1.14) | 1.06 (0.97-1.17) | 1.02 (0.95-1.09) | 1.01 (0.94-1.09) | 0.99 (0.93-1.07) | 1.05 (0.98-1.13) | **1.44 (1.32-1.56)** |

Results are HRs (95%CIs) per one SD increase of each MVX component, adjusted for age, sex, ethnicity, education level, TDI, smoking status, drinking status, diet, sleep duration, physical activity, parental history of CMM, hypertension, dyslipidemia, BMI, and HbA1c. **Bold** indicates statistical significance. HR, hazard ratio; CI, confidence interval; SD, standard deviation; FCMD, first cardiometabolic disease; CMM, cardiometabolic multimorbidity; GlycA, glycoprotein acetyls; S-HDLP, small high-density lipoprotein particles; IVX, inflammation vulnerability index; MMX, metabolic malnutrition index; TDI, Townsend deprivation index; BMI, body mass index.

**Table S11.** Hazard ratios (95%CIs) of MVX adjusted for regression dilution ratio.

|  | **Original HRs (95% CIs)** | **RDR-adjusted HRs (95%CIs)** |
| --- | --- | --- |
| **Incident CMDs** |  |  |
| CHD | **1.11 (1.09-1.13)** | **1.19 (1.16-1.23)** |
| Stroke | **1.04 (1.01-1.07)** | **1.07 (1.01-1.12)** |
| T2DM | **1.09 (1.07-1.12)** | **1.16 (1.11-1.20)** |
| **CMM status** |  |  |
| FCMD | **1.09 (1.08-1.10)** | **1.15 (1.13-1.18)** |
| CMM | **1.11 (1.07-1.15)** | **1.19 (1.11-1.27)** |
| **All-cause death** | **1.12 (1.10-1.14)** | **1.21 (1.17-1.25)** |
| **CMM transitions** |  |  |
| Baseline → FCMD | **1.09 (1.08-1.10)** | **1.15 (1.13-1.18)** |
| FCMD → CMM | **1.11 (1.08-1.14)** | **1.19 (1.14-1.24)** |
| Baseline → Death | **1.05 (1.01-1.10)** | **1.09 (1.02-1.16)** |
| FCMD → Death | **1.10 (1.06-1.13)** | **1.16 (1.10-1.23)** |
| CMM → Death | **1.17 (1.08-1.26)** | **1.29 (1.13-1.48)** |

Results are hazard ratios (95% confidence intervals) for MVX per 1 SD increase in the fully-adjusted model. **Bold** indicates statistical significance. CMD, cardiometabolic diseases; CHD, coronary heart disease; CI, confidence intervals; T2DM, type 2 diabetes mellitus; FCMD, first cardiometabolic disease; CMM, cardiometabolic multimorbidity; HR, hazard ratio; MVX, metabolic vulnerability index; SD, standard deviation.

**Table S12.** Hazard ratios (95%CIs) of MVX for each transition excluding participants who entered different states on the same date (*n* = 1,251).

| **Transitions** | **Event *n*** | **MVX, per one SD increase** |
| --- | --- | --- |
| **Baseline → FCMD** | 26,544 | **1.09 (1.07-1.10)** |
| **FCMD → CMM** | 9,220 | **1.06 (1.01-1.11)** |
| **Baseline → Death** | 2,847 | **1.11 (1.08-1.14)** |
| **FCMD → Death** | 3,145 | **1.07 (1.03-1.12)** |
| **CMM → Death** | 595 | **1.17 (1.07-1.28)** |

Results are hazard ratios (95% confidence intervals). **Bold** indicates statistical significance. FCMD, first cardiometabolic disease; CMM, cardiometabolic multimorbidity (defined as co-existence of ≥2 cardiometabolic diseases); MVX, metabolic vulnerability index; SD, standard deviation.

**Table S13.** Hazard ratios (95%CIs) of MVX for each transition when calculating the entering date of the prior state using median transition interval time.

| **Transitions** | **Event *n*** | **MVX, per one SD increase** |
| --- | --- | --- |
| **Baseline → FCMD** | 27,770 | **1.08 (1.07-1.10)** |
| **Baseline → Death** | 9,244 | **1.10 (1.07-1.12)** |
| **FCMD → CMM** | 2,970 | 1.04 (1.00-1.08) |
| **FCMD → Death** | 4,238 | **1.07 (1.03-1.11)** |
| **CMM → Death** | 729 | **1.13 (1.04-1.23)** |

Results are hazard ratios (95% confidence intervals). **Bold** indicates statistical significance. The median transition time was 1.97 years from FCMD to CMM, 0.31 years from FCMD to death, and 0.25 years from CMM to death. FCMD, first cardiometabolic disease; CMM, cardiometabolic multimorbidity (defined as co-existence of ≥2 cardiometabolic diseases); MVX, metabolic vulnerability index; SD, standard deviation.

**Table S14.** Hazard ratios (95%CIs) of MVX for each transition after adding another transition from the baseline directly to CMM.

| **Transitions** | **Event *n*** | **MVX, per one SD increase** |
| --- | --- | --- |
| **Baseline → FCMD** | 27,204 | **1.09 (1.07-1.10)** |
| **Baseline → CMM** | 601 | **1.11 (1.01-1.21)** |
| **Baseline → Death** | 9,220 | **1.11 (1.08-1.14)** |
| **FCMD → CMM** | 2,405 | **1.05 (1.01-1.10)** |
| **FCMD → Death** | 4,237 | **1.10 (1.06-1.13)** |
| **CMM → Death** | 754 | **1.19 (1.10-1.28)** |

Results are hazard ratios (95% confidence intervals). **Bold** indicates statistical significance. FCMD, first cardiometabolic disease; CMM, cardiometabolic multimorbidity (defined as co-existence of ≥2 cardiometabolic diseases); MVX, metabolic vulnerability index; SD, standard deviation.

**Table S15.** Hazard ratios (95%CIs) of MVX for CMDs, CMM, mortality, and each transition after redefining T2DM as ICD-10 code E11 (specified T2DM).

| **Transitions** | **Event *n*** | **MVX, per one SD increase** |
| --- | --- | --- |
| **Incident CMDs** |  |  |
| CHD | 16,615 | **1.11 (1.09-1.13)** |
| Stroke | 5,143 | **1.04 (1.01-1.07)** |
| T2DM | 1,407 | **1.07 (1.01-1.13)** |
| **CMM status** |  |  |
| FCMD | 21,897 | **1.09 (1.08-1.11)** |
| CMM | 1,246 | **1.11 (1.05-1.19)** |
| **All-cause death** | 14,211 | **1.12 (1.10-1.14)** |
| **CMM transitions** |  |  |
| Baseline → FCMD | 21,897 | **1.09 (1.08-1.11)** |
| Baseline → Death | 9,826 | **1.12 (1.09-1.14)** |
| FCMD → CMM | 1,246 | 1.04 (0.97-1.10) |
| FCMD → Death | 3,946 | **1.07 (1.04-1.11)** |
| CMM → Death | 439 | **1.24 (1.12-1.38)** |

Results are hazard ratios (95% confidence intervals). **Bold** indicates statistical significance. CHD, coronary heart disease; T2DM, type 2 diabetes mellitus; FCMD, first cardiometabolic disease; CMM, cardiometabolic multimorbidity (defined as co-existence of ≥2 cardiometabolic diseases); MVX, metabolic vulnerability index; SD, standard deviation.

**Table S16.** Hazard ratios (95%CIs) of MVX for CMDs, CMM, mortality, and each transition excluding participants with events occurring in the first two years of follow-up (*n* = 2,923).

| **Transitions** | **Event *n*** | **MVX, per one SD increase** |
| --- | --- | --- |
| **Incident CMDs** |  |  |
| CHD | 13,863 | **1.01 (1.08-1.12)** |
| Stroke | 4,436 | **1.04 (1.00-1.07)** |
| T2DM | 8,492 | **1.09 (1.07-1.12)** |
| **CMM status** |  |  |
| FCMD | 24,192 | **1.09 (1.07-1.10)** |
| CMM | 2,463 | **1.10 (1.05-1.15)** |
| **All-cause death** | 12,111 | **1.10 (1.08-1.13)** |
| **CMM transitions** |  |  |
| Baseline → FCMD | 24,192 | **1.09 (1.07-1.10)** |
| Baseline → Death | 8,758 | **1.10 (1.07-1.12)** |
| FCMD → CMM | 2,463 | 1.04 (0.99-1.10) |
| FCMD → Death | 2,862 | **1.08 (1.03-1.12)** |
| CMM → Death | 491 | **1.19 (1.07-1.32)** |

Results are hazard ratios (95% confidence intervals). **Bold** indicates statistical significance. CMD, cardiometabolic diseases; CHD, coronary heart disease; T2DM, type 2 diabetes mellitus; FCMD, first cardiometabolic disease; CMM, cardiometabolic multimorbidity (defined as co-existence of ≥2 cardiometabolic diseases); MVX, metabolic vulnerability index; SD, standard deviation.

**Table S17.** Hazard ratios (95%CIs) of MVX for CMDs, CMM, mortality, and each transition additionally adjusting for anti-hypertensive medication, cholesterol-lowering medication, albumin, and CRP.

| **Transitions** | **Event *n*** | **MVX, per one SD increase** |
| --- | --- | --- |
| **Incident CMDs** |  |  |
| CHD | 16,615 | **1.09 (1.07-1.11)** |
| Stroke | 51143 | 1.01 (0.98-1.04) |
| T2DM | 91210 | **1.07 (1.04-1.09)** |
| **CMM status** |  |  |
| FCMD | 27,805 | **1.06 (1.05-1.08)** |
| CMM | 3,006 | **1.06 (1.02-1.10)** |
| **All-cause death** | 14,211 | **1.06 (1.04-1.08)** |
| **CMM transitions** |  |  |
| Baseline → FCMD | 27,805 | **1.08 (1.07-1.09)** |
| FCMD → CMM | 3,006 | 1.03 (0.99-1.07) |
| Baseline → Death | 9,220 | **1.09 (1.06-1.12)** |
| FCMD → Death | 4,237 | **1.05 (1.01-1.08)** |
| CMM → Death | 754 | 1.08 (1.00-1.17) |

Results are hazard ratios (95% confidence intervals). **Bold** indicates statistical significance. CRP, C-reactive protein; CMD, cardiometabolic diseases; CHD, coronary heart disease; T2DM, type 2 diabetes mellitus; FCMD, first cardiometabolic disease; CMM, cardiometabolic multimorbidity (defined as co-existence of ≥2 cardiometabolic diseases); MVX, metabolic vulnerability index; SD, standard deviation.

**Table S18.** Hazard ratios (95%CIs) of MVX for CMDs, CMM, mortality, and each transition excluding participants with missing values in covariates (*n* = 73,741).

| **Transitions** | **Event *n*** | **MVX, per one SD increase** |
| --- | --- | --- |
| **Incident CMDs** |  |  |
| CHD | 10,295 | **1.11 (1.08-1.13)** |
| Stroke | 3,167 | 1.04 (1.00-1.08) |
| T2DM | 5,326 | **1.09 (1.06-1.12)** |
| **CMM status** |  |  |
| FCMD | 16,988 | **1.09 (1.07-1.10)** |
| CMM | 1,714 | **1.12 (1.07-1.18)** |
| **All-cause death** | 8,405 | **1.10 (1.07-1.13)** |
| **CMM transitions** |  |  |
| Baseline → FCMD | 16,988 | **1.09 (1.07-1.11)** |
| Baseline → Death | 5,544 | **1.08 (1.05-1.11)** |
| FCMD → CMM | 1,714 | **1.07 (1.02-1.13)** |
| FCMD → Death | 2,433 | **1.08 (1.03-1.13)** |
| CMM → Death | 428 | **1.15 (1.03-1.28)** |

Results are hazard ratios (95% confidence intervals). **Bold** indicates statistical significance. CMD, cardiometabolic diseases; CHD, coronary heart disease; T2DM, type 2 diabetes mellitus; FCMD, first cardiometabolic disease; CMM, cardiometabolic multimorbidity; MVX, metabolic vulnerability index; SD, standard deviation.

**Table S19.** Hazard ratios (95%CIs) of MVX for CMDs, CMM, mortality, and each transition using data imputed with multiple imputation by chained equations.

| **Transitions** | **Event *n*** | **MVX, per one SD increase** |
| --- | --- | --- |
| **Incident CMDs** |  |  |
| CHD | 16,615 | **1.11 (1.09-1.13)** |
| Stroke | 5,143 | **1.04 (1.00-1.07)** |
| T2DM | 9,120 | **1.08 (1.06-1.11)** |
| **CMM status** |  |  |
| FCMD | 27,805 | **1.08 (1.07-1.10)** |
| CMM | 3,006 | **1.10 (1.06-1.14)** |
| **All-cause death** | 14,211 | **1.12 (1.10-1.14)** |
| **CMM transitions** |  |  |
| Baseline → FCMD | 27,805 | **1.08 (1.07-1.10)** |
| FCMD → CMM | 3,006 | **1.05 (1.01-1.09)** |
| Baseline → Death | 9,220 | **1.11 (1.08-1.13)** |
| FCMD → Death | 4,237 | **1.10 (1.06-1.13)** |
| CMM → Death | 754 | **1.17 (1.08-1.27)** |

Results are hazard ratios (95% confidence intervals). **Bold** indicates statistical significance. CMD, cardiometabolic diseases; CHD, coronary heart disease; T2DM, type 2 diabetes mellitus; FCMD, first cardiometabolic disease; CMM, cardiometabolic multimorbidity; MVX, metabolic vulnerability index; SD, standard deviation.

**Table S20.** Hazard ratios (95%CIs) of MVX for CMDs, CMM, mortality, and each transition not excluding participants with cancer at baseline (*n* = 24,893).

| **Transitions** | **Event *n*** | **MVX, per one SD increase** |
| --- | --- | --- |
| **Incident CMDs** |  |  |
| CHD | 17,495 | **1.11 (1.09-1.12)** |
| Stroke | 5,581 | **1.05 (1.02-1.08)** |
| T2DM | 10,083 | **1.09 (1.07-1.12)** |
| **CMM status** |  |  |
| FCMD | 29,732 | **1.09 (1.07-1.10)** |
| CMM | 3,253 | **1.11 (1.07-1.16)** |
| **All-cause death** | 15,971 | **1.15 (1.13-1.17)** |
| **CMM transitions** |  |  |
| Baseline → FCMD | 29,732 | **1.09 (1.07-1.10)** |
| FCMD → CMM | 3,253 | **1.07 (1.02-1.11)** |
| Baseline → Death | 11,508 | **1.15 (1.13-1.18)** |
| FCMD → Death | 3,737 | **1.09 (1.05-1.13)** |
| CMM → Death | 726 | **1.15 (1.06-1.25)** |

Results are hazard ratios (95% confidence intervals). **Bold** indicates statistical significance. CMD, cardiometabolic diseases; CHD, coronary heart disease; T2DM, type 2 diabetes mellitus; FCMD, first cardiometabolic disease; CMM, cardiometabolic multimorbidity; MVX, metabolic vulnerability index; SD, standard deviation.

**Table S21.** Hazard ratios (95%CIs) of MVX and its components for each transition not excluding participants with CMDs at baseline (*n* = 29,893).

| **Transitions** | **Event *n*** | **MVX, per one SD increase** |
| --- | --- | --- |
| **Baseline → FCMD** | 29,607 | **1.12 (1.11-1.14)** |
| **Baseline → Death** | 9,308 | **1.11 (1.09-1.14)** |
| **FCMD → CMM** | 8,529 | **1.10 (1.08-1.13)** |
| **FCMD → Death** | 7,102 | **1.13 (1.10-1.16)** |
| **CMM → Death** | 3,672 | **1.12 (1.08-1.17)** |

Results are hazard ratios (95% confidence intervals). **Bold** indicates statistical significance. Participants with CMDs at baseline (*n* = 29,893) were assigned to FCMD or CMM state according to their CMD status. CMD, cardiometabolic diseases; FCMD, first cardiometabolic disease; CMM, cardiometabolic multimorbidity; MVX, metabolic vulnerability index; SD, standard deviation.

**Table S22.** Hazard ratios (95%CIs) of MVX for CMDs, CMM, mortality, and each transition additionally adjusted for statins use.

|  | **Event *n*** | **MVX, per one SD increase** |
| --- | --- | --- |
| **Incident CMDs** |  |  |
| CHD | 16,615 | **1.11 (1.09-1.13)** |
| Stroke | 5,143 | **1.04 (1.01-1.07)** |
| T2DM | 9,120 | **1.10 (1.07-1.12)** |
| **CMM status** |  |  |
| FCMD | 27,805 | **1.09 (1.08-1.11)** |
| CMM | 3,006 | **1.11 (1.07-1.16)** |
| **All-cause death** | 14,211 | **1.12 (1.10-1.14)** |
| **CMM transitions** |  |  |
| Baseline → FCMD | 27,805 | **1.09 (1.08-1.11)** |
| FCMD → CMM | 3,006 | **1.05 (1.01-1.09)** |
| Baseline → Death | 9,220 | **1.11 (1.08-1.14)** |
| FCMD → Death | 4,237 | **1.09 (1.05-1.13)** |
| CMM → Death | 754 | **1.16 (1.07-1.26)** |

Results are hazard ratios (95% confidence intervals). **Bold** indicates statistical significance. CMD, cardiometabolic diseases; CHD, coronary heart disease; T2DM, type 2 diabetes mellitus; FCMD, first cardiometabolic disease; CMM, cardiometabolic multimorbidity; MVX, metabolic vulnerability index; SD, standard deviation.

**Table S23.** Results of stratified analysis by the use of statins.

| **Outcome** | **Subgroup** | **Event *n* (%)** | **HR (95%CI)** |
| --- | --- | --- | --- |
| CHD | Statin users | 3,508 (14.5%) | 1.04 (1.00-1.08) |
|  | Non-statin users | 13,107 (6.7%) | **1.13 (1.10-1.15)** |
| Stroke | Statin users | 940 (3.9%) | 1.01 (0.94-1.08) |
|  | Non-statin users | 4,203 (2.2%) | **1.04 (1.01-1.08)** |
| T2DM | Statin users | 2,270 (9.4%) | **1.08 (1.03-1.13)** |
|  | Non-statin users | 6,940 (3.6%) | **1.09 (1.06-1.12)** |
| FCMD | Statin users | 5,873 (24.3%) | **1.05 (1.02-1.08)** |
|  | Non-statin users | 21,932 (11.2%) | **1.10 (1.08-1.11)** |
| CMM | Statin users | 797 (3.3%) | 1.07 (0.99-1.15) |
|  | Non-statin users | 2,209 (1.1%) | **1.12 (1.07-1.17)** |
| Death | Statin users | 2,341 (9.7%) | **1.14 (1.09-1.19)** |
|  | Non-statin users | 11,870 (6.1%) | **1.12 (1.09-1.14)** |

Hazard ratios (95% confidence intervals) are for MVX, per 1 SD increase. **Bold** indicates statistical significance. CMD, cardiometabolic diseases; CHD, coronary heart disease; T2DM, type 2 diabetes mellitus; FCMD, first cardiometabolic disease; CMM, cardiometabolic multimorbidity; MVX, metabolic vulnerability index; SD, standard deviation.

**Table S24.** Hazard ratios (95%CIs) of MVX for CMDs, CMM, mortality, and each transition additionally adjusted for serum phosphate.

|  | **Event *n*** | **MVX, per one SD increase** |
| --- | --- | --- |
| **Incident CMDs** |  |  |
| CHD | 16,615 | **1.11 (1.09-1.13)** |
| Stroke | 5,143 | **1.04 (1.01-1.07)** |
| T2DM | 9,120 | **1.09 (1.07-1.12)** |
| **CMM status** |  |  |
| FCMD | 27,805 | **1.09 (1.08-1.10)** |
| CMM | 3,006 | **1.11 (1.07-1.15)** |
| **All-cause death** | 14,211 | **1.12 (1.10-1.14)** |
| **CMM transitions** |  |  |
| Baseline → FCMD | 27,805 | **1.09 (1.08-1.11)** |
| FCMD → CMM | 3,006 | **1.05 (1.01-1.09)** |
| Baseline → Death | 9,220 | **1.11 (1.08-1.14)** |
| FCMD → Death | 4,237 | **1.10 (1.06-1.13)** |
| CMM → Death | 754 | **1.15 (1.06-1.25)** |

Results are hazard ratios (95% confidence intervals). **Bold** indicates statistical significance. CMD, cardiometabolic diseases; CHD, coronary heart disease; T2DM, type 2 diabetes mellitus; FCMD, first cardiometabolic disease; CMM, cardiometabolic multimorbidity; MVX, metabolic vulnerability index; SD, standard deviation.

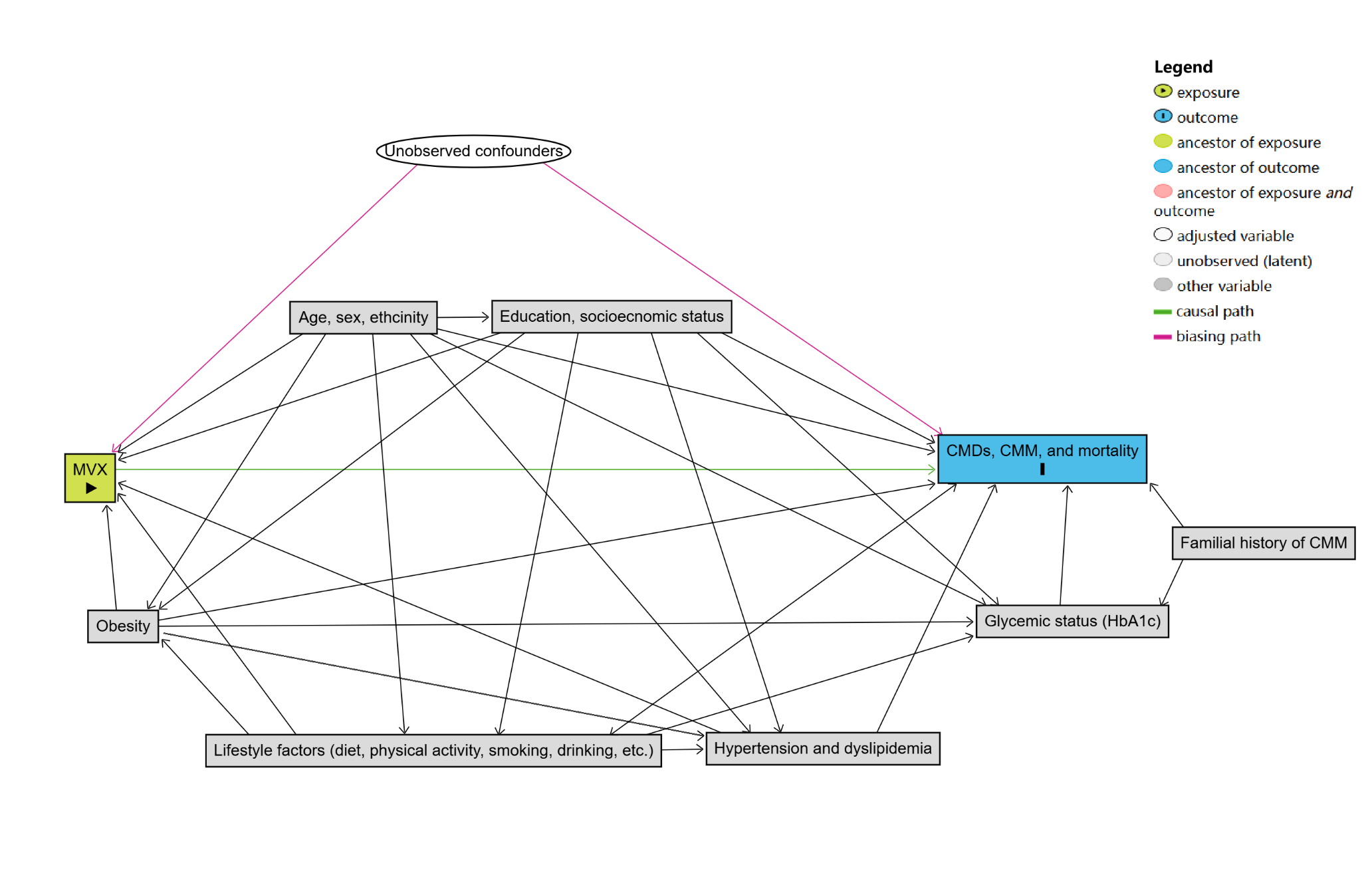

**Figure S1.** *A priori* defined directed acyclic graph for confounders in the MVX-CMDs/CMM association.

MVX, metabolic vulnerability index; CMD, cardiometabolic diseases; CMM, cardiometabolic multimorbidity; HbA1c, glycated hemoglobin A1c.

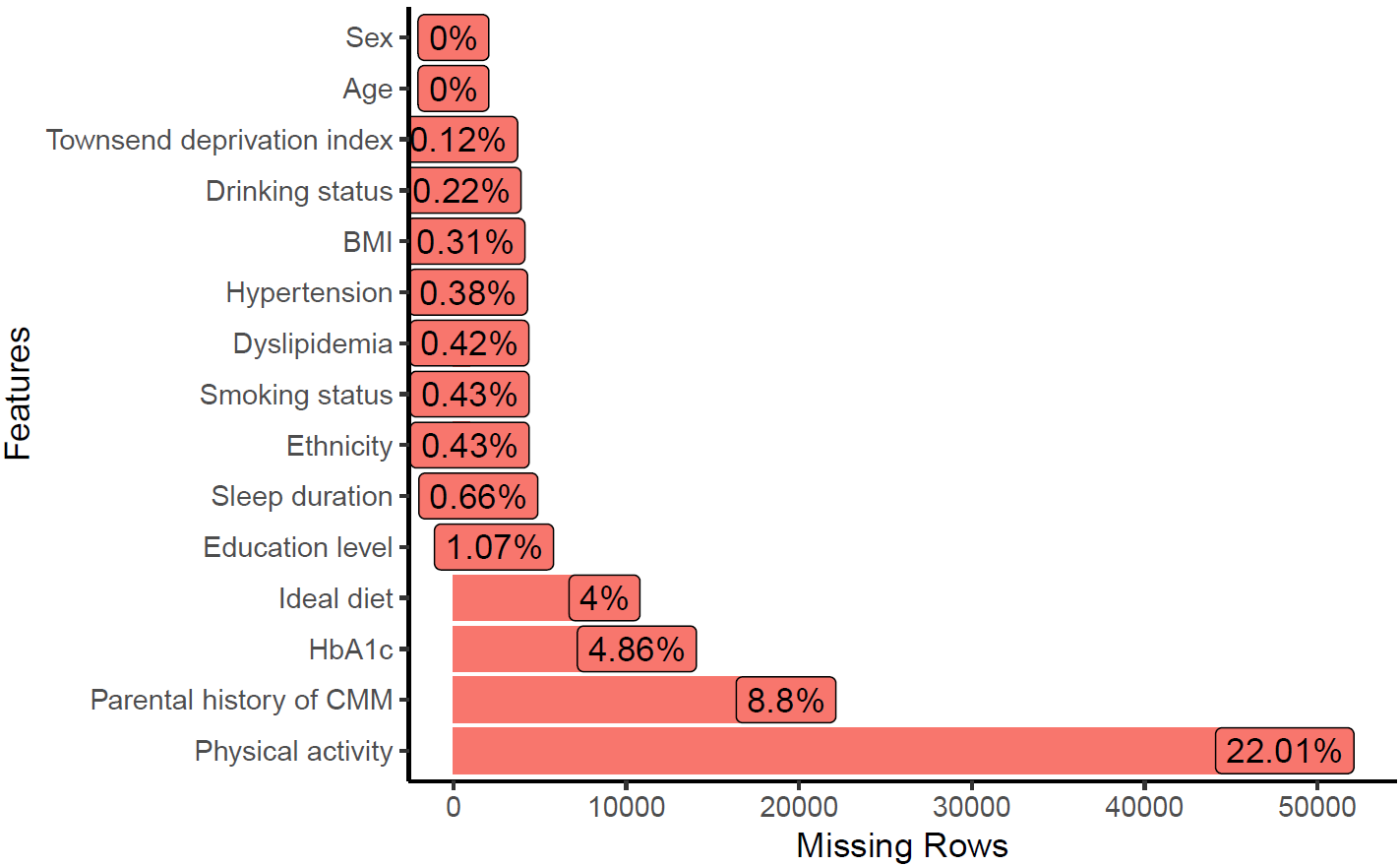

**Figure S2.** Numbers and percentages of missing values in covariates.

BMI, body mass index; HbA1c, glycated hemoglobin A1c; CMM, cardiometabolic multimorbidity.

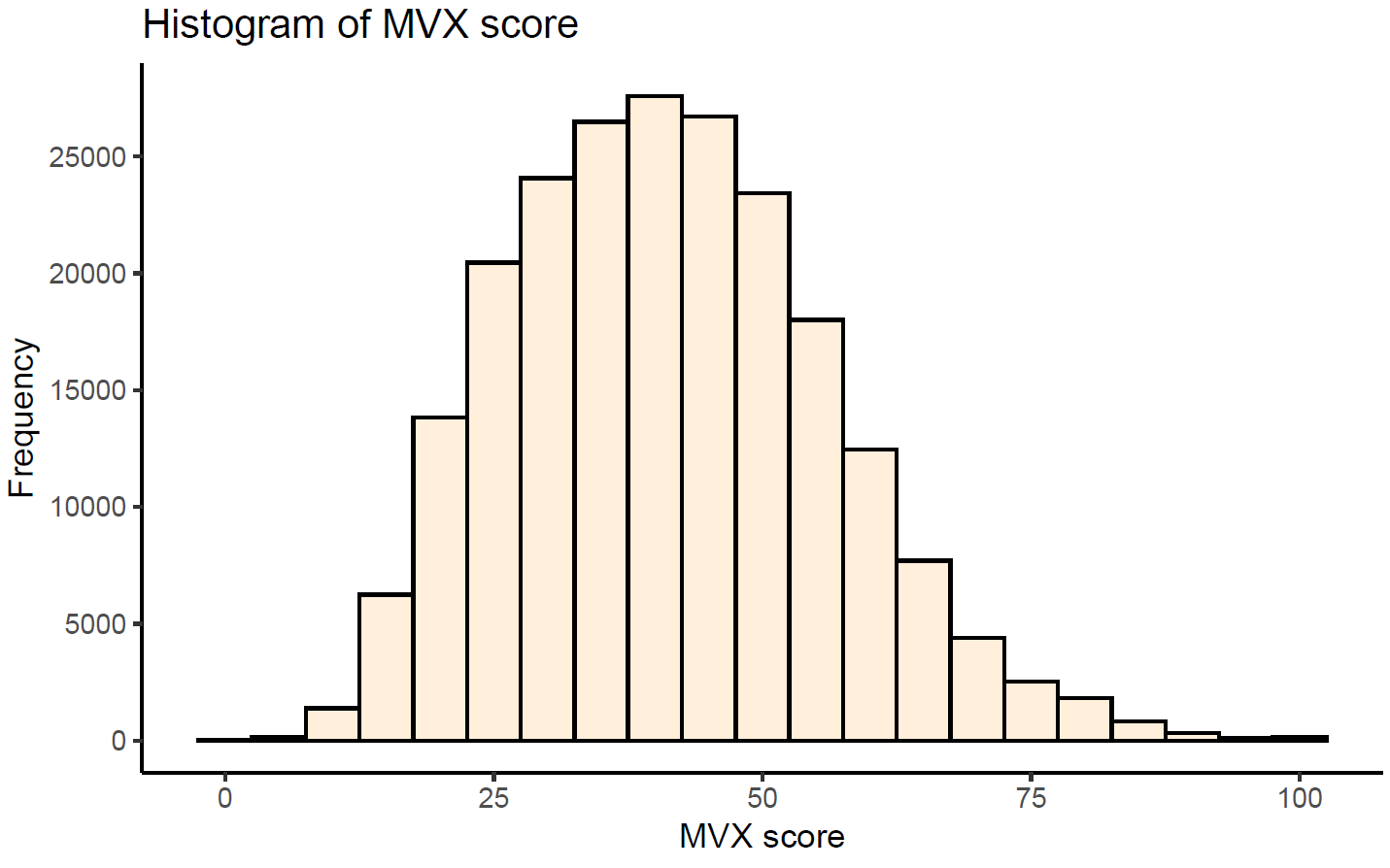

**Figure S3.** Histogram of MVX score in the UK Biobank cohort.

MVX, metabolic vulnerability index.

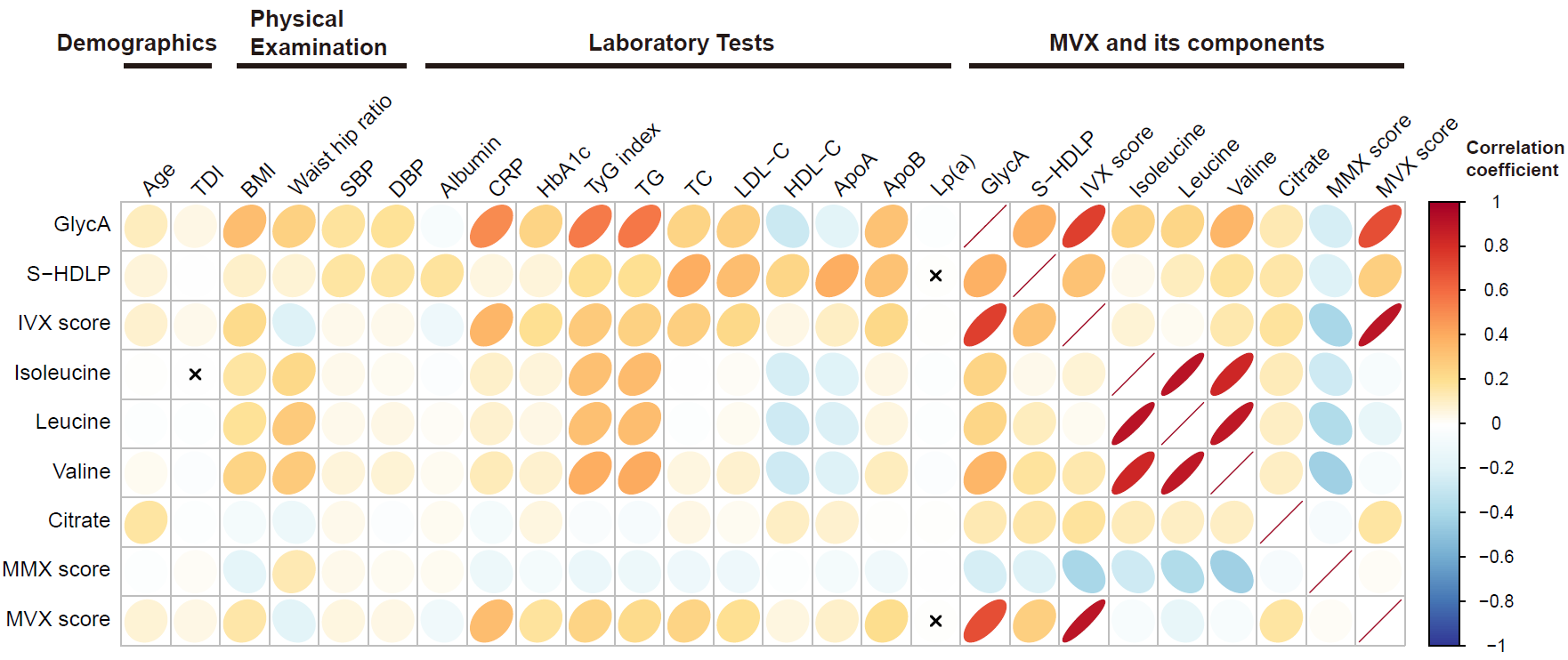

**Figure S4.** Results of the correlation analysis.

The color and orientation of the ellipses represents the correlation coefficient of the corresponding variables, and “×” represents insignificant correlations. Pearson or Spearman correlation analysis were used where appropriate. TDI, Townsend deprivation index; BMI, body mass index; SBP, systolic blood pressure; DBP, diastolic blood pressure; CRP, C-reactive protein; HbA1c, glycated hemoglobin A1c; TyG index, triglyceride-glucose index (represents insulin resistance); TG, triglycerides; TC, total cholesterol; LDL-C, low-density lipoprotein-cholesterol; HDL-C, high-density lipoprotein-cholesterol; ApoA, apolipoprotein A; ApoB, apolipoprotein B; Lp(a), lipoprotein(a).

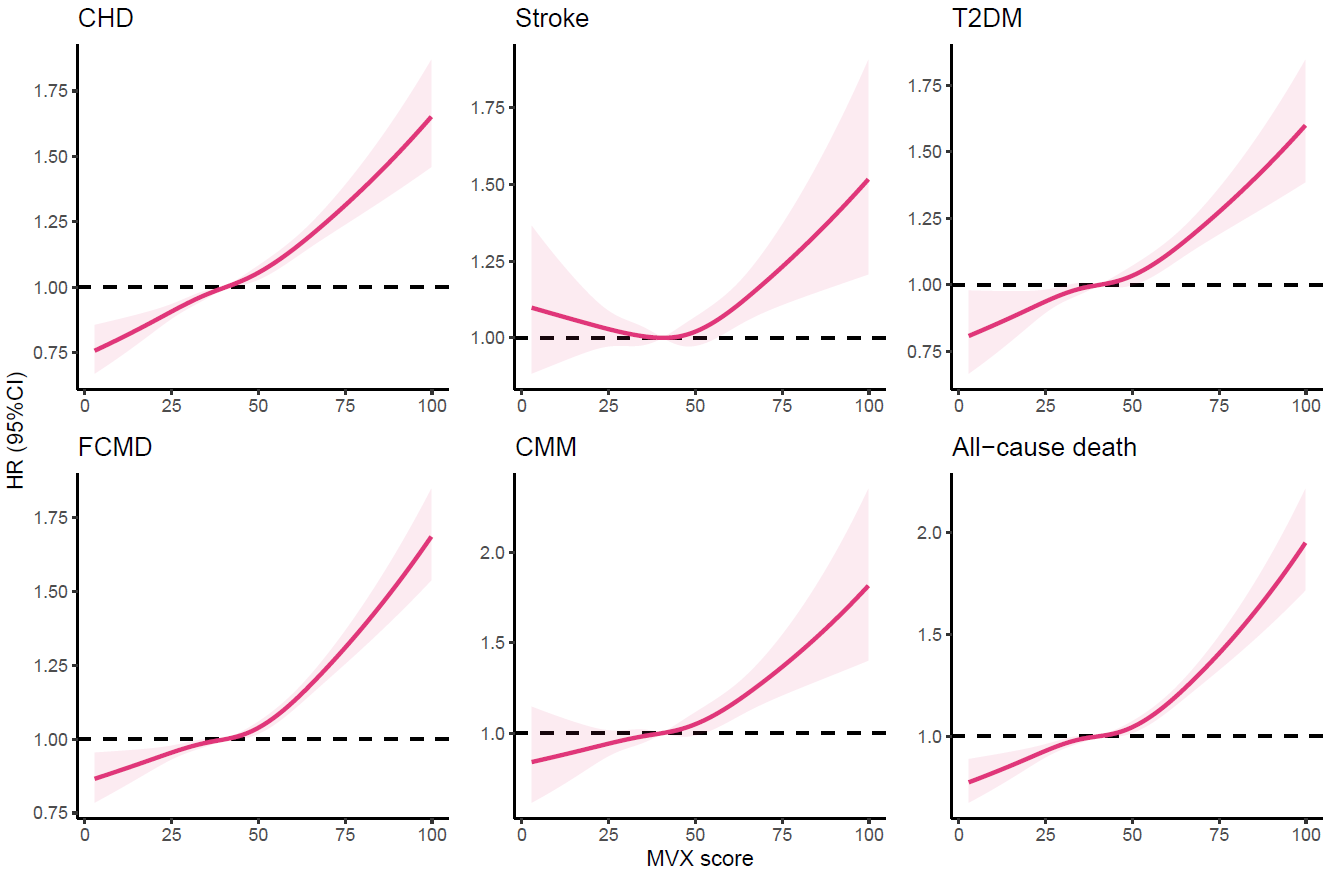

**Figure S5.** Restrictive cubic splines of MVX score for the sequential onset patterns of CMDs.

HR, hazard ratio; CI, confidence interval; CHD, coronary heart disease; T2DM, type 2 diabetes mellitus; FCMD, first cardiometabolic disease; CMM, cardiometabolic multimorbidity; MVX, metabolic vulnerability index

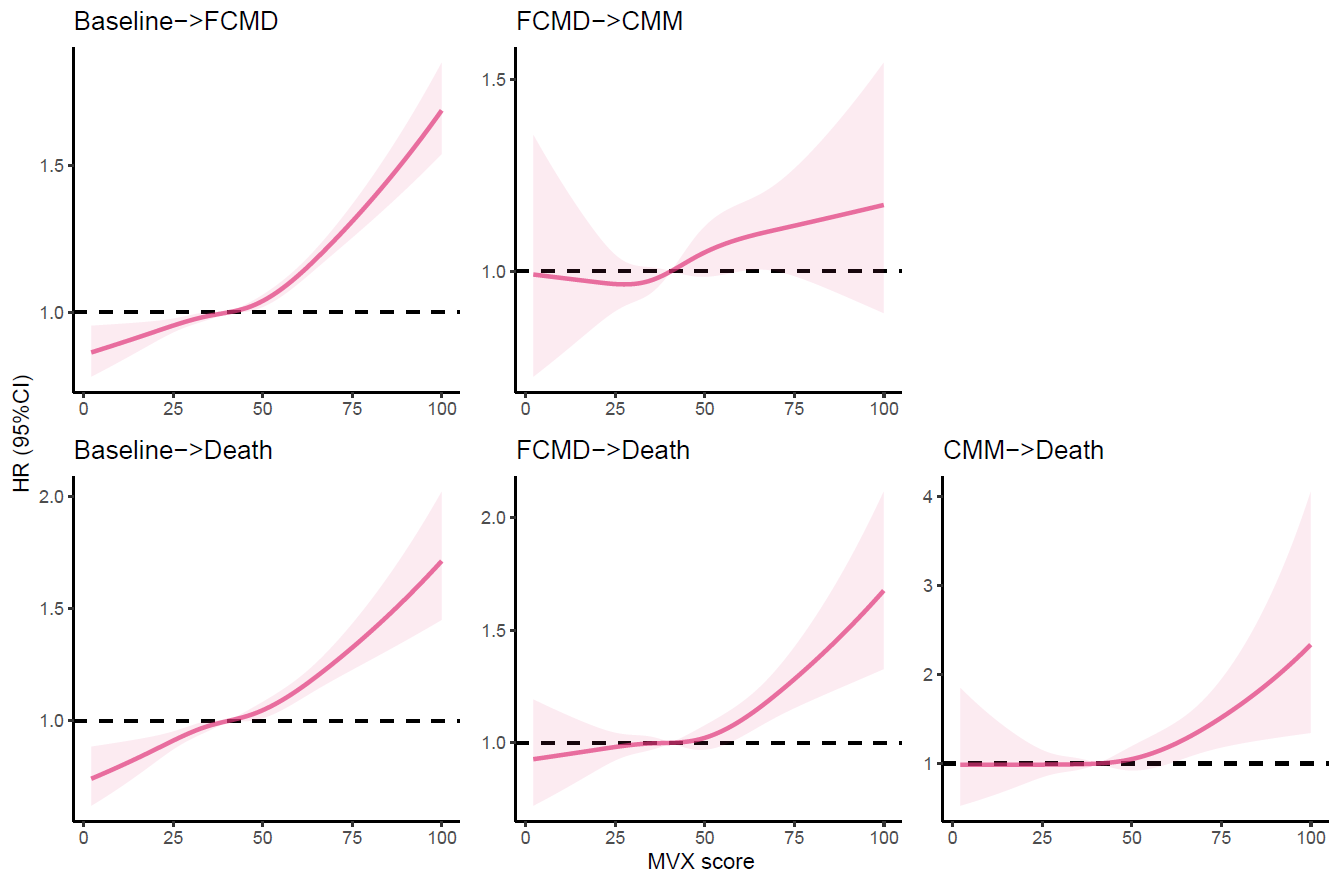

**Figure S6.** Restrictive cubic splines of MVX score for each transition in pattern A.

HR, hazard ratio; CI, confidence interval; FCMD, first cardiometabolic disease; CMM, cardiometabolic multimorbidity; MVX, metabolic vulnerability index

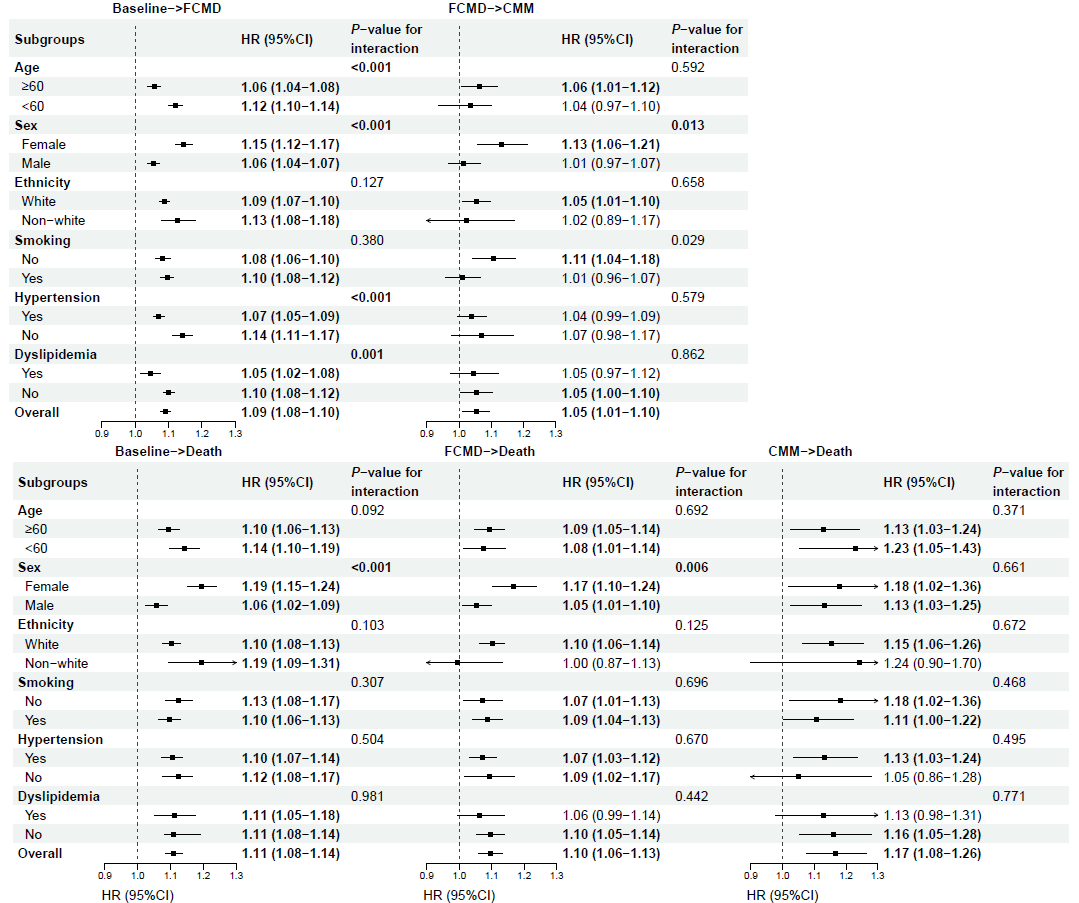

**Figure S7.** Hazard ratios (95% CIs) of MVX score for each transition in the stratified analysis.

Results are hazard ratios (95% CIs) per one SD increase of MVX. **Bold** indicates statistical significance. CI, confidence interval; FCMD, first cardiometabolic disease; CMM, cardiometabolic multimorbidity; MVX, metabolic vulnerability index; SD, standard deviation.
